# Novel Epistatic Interaction Between RBMS3 and CDKN2B-AS1 in Coronary Artery Disease Risk Identified by Machine Learning Tool VariantSpark

**DOI:** 10.1101/2025.10.19.25338331

**Authors:** Letitia M.F. Sng, Mitchell J. O’Brien, Brendan Hosking, Piotr Szul, Roc Reguant, Mythreye Venkatesan, Philip J. Freda, Zhiping Wang, Jason H. Moore, Anne H. Klein, Michael Kuiper, Angus Panagopoulos, Johan W. Verjans, Yatish Jain, Denis C. Bauer, Natalie A. Twine

## Abstract

**Background:** Genome-wide association studies (GWAS) of coronary artery disease (CAD), the leading cause of mortality and morbidity globally, have identified approximately 163 risk loci, yet only 40% of CAD genetic heritability can be explained. Non-additive genetic effects, like epistasis, likely contribute to CAD aetiology, but remain elusive due to limited data and insensitive algorithms.

**Results:** Using machine-learning (VariantSpark) followed by exhaustive epistasis search (BitEpi), we discovered an epistatic interaction for CAD between *RBMS3* and *CDKN2B-AS1* in the UK Biobank. We also observe this interaction in the independent All of Us cohort and provide a binding model using AlphaFold3. VariantSpark provides the needed sensitivity, identifying associated loci (e.g. *PMAIP1-MC4R* and *AAK1)* with 72% fewer samples than previous studies, to reduce the search space for systematic epistasis detection.

**Conclusions:** We provide *in silico* evidence of *RBMS3* as a novel CAD risk gene, acting in epistasis with the established *CDKN2B-AS1* 9p21.3 risk loci.

## Background

Coronary artery disease (CAD) is the leading cause of mortality and morbidity globally with a strong genetic component in its aetiology (1,2). Since the first genome-wide association studies (GWAS) in 2007 (3,4), approximately 897 independent loci have been associated to CAD, but only cumulatively explain 35-40% of CAD heritability on the liability scale (5,6). Furthermore, one study estimated a 34.3% heritability even with the inclusion of ultra-rare variants (7), suggesting other sources of the missing heritability observed.

Epistasis, the combinatorial effect of one or more genes which may act alongside or in absence of marginal effects, has been suggested to be part of the solution but has not yet been systematically studied. This is despite empirical evidence demonstrating its role in the genetic architecture of CAD (8–13). This absence of epistasis studies is because the combinatorial nature of epistasis analysis results in challenges for traditional parametric statistical methods such as logistic regression due to: (i) the curse of dimensionality where much larger sample sizes are required when searching for epistatic interactions than for additive effects (14); (ii) the need to specify interactions which is difficult due to the levels of *k* (i.e., 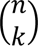) that need to be considered (15), and (iii) the computational demand and high multiple testing burden.

In this study, we address these challenges with an innovative two-stepped approach combining two established algorithms in a complementary way that provides higher sensitivity than previous GWAS approaches and evaluates non-linear epistatic interactions (16). First, VariantSpark (17), a cloud-based, scalable, machine learning (ML) GWAS tool is applied to search for putative interactions genome-wide. VariantSpark is based on a random forest framework using the Gini importance score to identify associations with both individual and interacting effects. This enables a purely data-driven and unbiased interaction discovery without requiring pre-specified models. VariantSpark effectively narrows the search space from 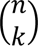 of possible interactions to the set of likely ones that can be evaluated systematically in the second step: BitEpi (18), a fast, exhaustive epistasis detection algorithm, is then used to mathematically test and annotate the exact epistatic interaction partners in the dataset.

By first filtering epistasis candidates with VariantSpark, we substantially reduce the multiple testing burden associated with such a search, making it feasible to analyse datasets large enough to observe the subtle effects of epistasis. We apply this approach to a CAD cohort within the UK Biobank (UKB) and TOPMed (TM) datasets to identify putative epistasis signals associated with CAD.

## Results

### VariantSpark Identifies Novel CAD Associations in a UK Biobank Cohort and Demonstrates Higher Sensitivity

VariantSpark was run on the imputed array genotype data from a CAD cohort within the UK Biobank (19) (UKB, n = 51,107) and validated in the Trans-Omics for Precision Medicine (TOPMed) (20) dataset (TM, n = 11,326). 115 single nucleotide polymorphisms (SNPs) significantly associated to CAD were identified in the UKB dataset (Supplementary Table 1, see file ‘Supplementary_Tables’), of which 99 (89.09%) were previously known and annotated in the Cardiovascular Disease Knowledge Portal (CVD KP) (21,22). This included well established CAD risk loci, namely 6q25.3 (SLC22A3), 6q26 (LPA), and 9p21.3 (CDKN2B-AS1) (Figure 1).

**Figure 1.**
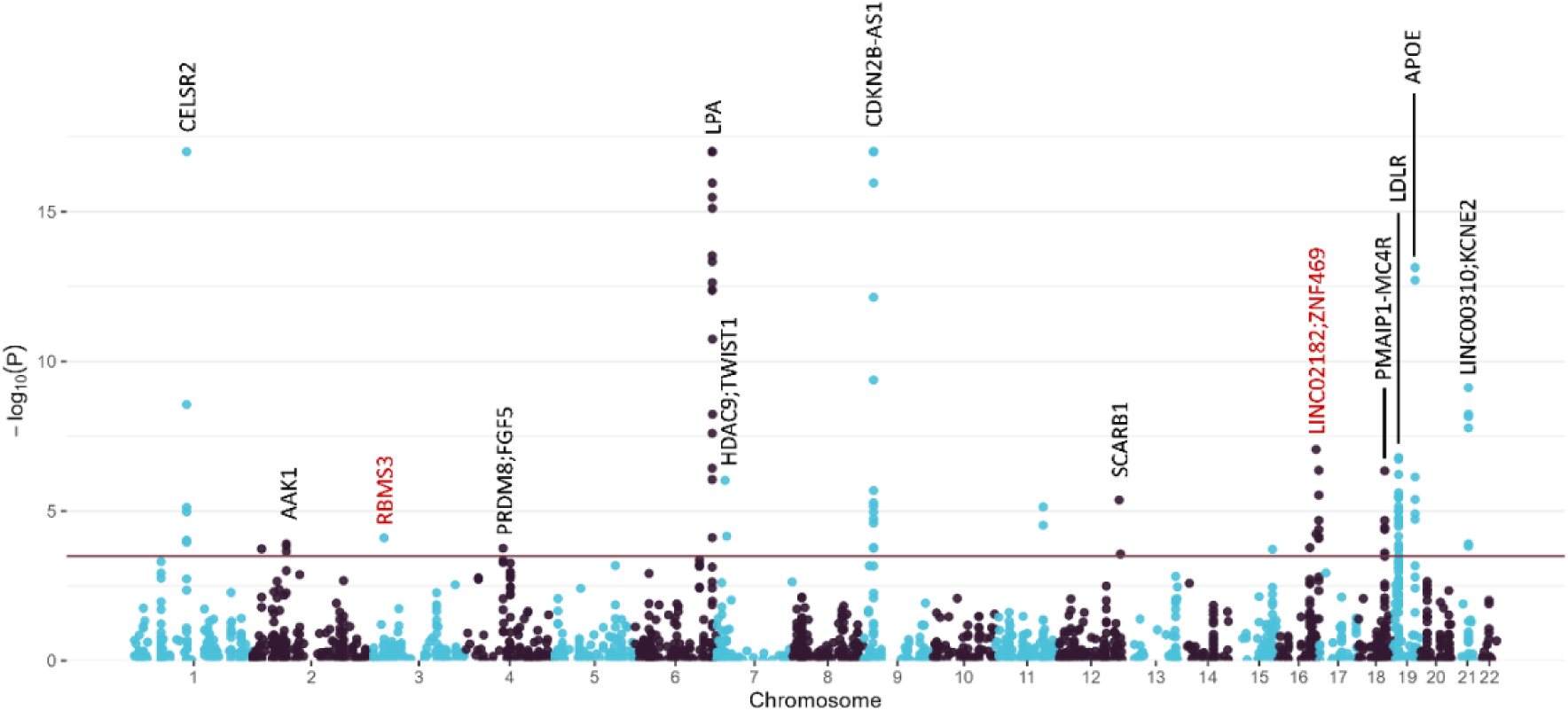
Manhattan plot of genome-wide association analysis using VariantSpark on UK Biobank CAD Cohort of 51,107 samples. The plot displays −log_10_(P) values calculated using VariantSpark’s RFlocalfdr implementation (*23*), with the horizontal line indicating the genome-wide significance threshold (localFDR < 5%). Notable loci are labelled, with established CAD-associated genes in black and novel associations highlighted in red (including *RBMS3*). The VariantSpark Manhattan plot appears sparser than traditional logistic regression plots because the RFlocalFDR approach assigns p-values to SNPs meeting specific criteria only. Key significant associations are visible at chromosomes 1 (*CELSR2*), 6 (*LPA*), 9 (*CDKN2B-AS1*), and 19 (*APOE*), with novel findings at chormosomes 3 (*RBMS3*), 16 (*WWP2*, *ZNF469*), and others. SNPs have not been pruned for linkage disequilibrium.

The 115 associations were clumped to 24 independent loci, and 8 loci were not previously associated to CAD in the CVD KP and/or GWAS Catalog (Table 1). These 8 loci may not have been picked up by traditional GWAS approaches as their individual effects are not large. In contrast, VariantSpark also includes interaction effects and hence increases sensitivity especially when boosted through its purpose-build local false discovery rate approach (23) increasing the chance of finding biologically meaningful effects. Indeed, two lead SNPs on chromosome 16 have evidence of association to CAD in previous studies: *rs*10852491 (*WWP2*) is in moderate LD (R^2^ = 0.397) with CAD-associated SNPs (6,24) and *rs*72803480 (*ZNF469*) has been marginally associated with coronary atherosclerosis on the PheWeb (P = 1.8 × 10^-6^) (25). At the gene level, *CDH13* (5,26) and *AAK1* (5,6) have been significantly associated to CAD in previous GWAS although the identified SNPs in these studies are not in high LD (R^2^ > 0.2) with those identified in our study.

**Table 1.** Significant Lead SNPs from VariantSpark GWAS with UK BioBank Cohort.

| Chr:Pos | R | A | RSID | Variant Function | Closest Gene(s) | GI | GI (P) | Known |
| --- | --- | --- | --- | --- | --- | --- | --- | --- |
| 1:56914738 | T | C | rs6684929 | intergenic | <u>LINC01767</u>;PLPP3 | 0.06 | 4.9E-4 | TRUE |
| 1:109817590 | G | T | rs12740374 | UTR3 | CELSR2* | 4.27 | 0 | TRUE |
| 2:18684371 | A | C | rs6743746 | intergenic | KCNS3;<u>RDH14</u> | 0.07 | 1.85E-4 | FALSE |
| 2:69839825 | T | A | rs2312215 | intronic | AAK1* | 0.07 | 1.27E-4 | FALSE |
| 3:29357538 | G | A | rs74467064 | intronic | RBMS3 | 0.07 | 7.93E-5 | FALSE |
| 4:81182554 | T | C | rs12509595 | intergenic | PRDM8;<u>FGF5</u> | 0.07 | 1.77E-4 | TRUE |
| 6:160759840 | C | T | rs386453 | intergenic | SLC22A2;<u>SLC22A3</u>* | 0.16 | 3.73E-7 | TRUE |
| 6:160922870 | A | G | rs117733303 | ncRNA (intronic) | LPAL2 | 1.22 | 0 | TRUE |
| 6:160985526 | G | A | rs118039278 | intronic | LPA* | 10.75 | 0 | TRUE |
| 7:22300682 | T | C | rs77924461 | intronic | RAPGEF5 | 0.08 | 6.97E-5 | FALSE |
| 7:19049388 | G | A | rs2107595 | intergenic | <u>HDAC9</u>;TWIST1* | 0.14 | 9.48E-7 | TRUE |
| 9:22085598 | T | C | rs1970112 | ncRNA (intronic) | CDKN2B-AS1* | 2.10 | 0 | TRUE |
| 11:100596548 | C | T | rs11224412 | intronic | ARHGAP42* | 0.11 | 7.32E-6 | TRUE |
| 12:125316055 | A | C | rs11057840 | intronic | SCARB1 | 0.12 | 4.31E-6 | TRUE |
| 12:128065186 | G | A | rs12367091 | intergenic | LINC02375;<u>LINC02411</u> | 0.07 | 2.75E-4 | FALSE |
| 15:91425232 | C | T | rs6227 | UTR3 | FURIN* | 0.07 | 1.9E-4 | TRUE |
| 16:69928846 | T | C | rs10852461 | intronic | WWP2 | 0.07 | 1.68E-4 | FALSE |
| 16:82732474 | G | A | rs77684808 | intronic | CDH13 | 0.19 | 8.79E-8 | FALSE |
| 16:88382789 | C | T | rs72803480 | intergenic | LINC02182;<u>ZNF469</u> | 0.16 | 4.38E-7 | FALSE |
| 18:57937459 | A | G | rs718475 | intergenic | PMAIP1;<u>MC4R</u> | 0.16 | 4.57E-7 | TRUE |
| 19:11206729 | T | C | rs17242381 | intronic | LDLR | 0.17 | 1.64E-7 | TRUE |
| 19:45412079 | C | T | rs7412 | exonic | APOE | 0.69 | 7.37E-14 | TRUE |
| 19:45415713 | G | A | rs10414043 | intergenic | APOE*;<u>APOC1</u> | 0.15 | 7.32E-7 | TRUE |
| 21:35599128 | C | T | rs9982601 | intergenic | <u>LINC00310</u>*;KCNE2* | 0.31 | 7.63E-10 | TRUE |

Ordered by chromosomal location. GI = Gini Importance score from VariantSpark, GI (P) = VariantSpark P-value. Known SNPs were previously associated to CAD in CVD KP and/or GWAS Catalog. Underlined genes indicate closest gene, \* indicates eQTL on STARNET.

Applying VariantSpark to the independent but smaller TM dataset replicated 14 SNPs at genome-wide significance (*P* < 5 × 10^-8^) (Supplementary Table 2, see file ‘Supplementary_Tables’), mapping to two established CAD risk loci: *LPA* and *CDKN2B-AS1*. In comparison, a previous study replicated 14 SNPs between UKB and TM but below genome-wide significance levels (*P* < 0.001) (27).

At the gene-level, *CDH13* replicated across both UKB and TM cohorts, however, the identified SNPs from each cohort were not in LD. This is likely due to the heterogeneity in the TM cohort, which was formed by harmonising multiple independent studies (e.g., Framingham and the Women’s Health Study). As a result, a clearly defined CAD phenotype was lacking, unlike the UKB dataset where the availability and use of ICD codes enabled a more precise CAD definition.

To compare VariantSpark’s performance against a more traditional GWAS approach, we applied PLINK’s logistic regression (28) on the same UKB and TM cohorts. For UKB, PLINK identified 117 significant SNPs, clumping to 10 risk loci. VariantSpark found 9 of the 10 PLINK identified loci except *TRIB1* on chromosome 8 (Supplementary Table 3, see file ‘Supplementary_Tables’).

VariantSpark assigned a *p*-value of 0.30 to *TRIB1*, likely because it is an independent association and does not reach significance amongst the other features. In the TM cohort, PLINK only identified 5 significant SNPs, all mapping to one locus, *CDKN2B-AS1* (Supplementary Table 4, see file ‘Supplementary_Tables’). In contrast, VariantSpark also identified the well-known *LPA* locus, which in the PLINK analysis failed genome-wide significance for TM (5 × 10^-8^ < *P* < 1 ×10^-5^), confirming VariantSpark’s higher sensitivity, also observed in Alzheimer’s (16).

### The novel CAD-risk gene, *RBMS3*, is likely involved in CAD pathogenesis through interaction with *CDKN2B-AS1*

Next, we conducted an epistasis search using BitEpi on the larger UKB cohort with the well-defined CAD phenotype. Specifically, BitEpi exhaustively searched for pairwise interactions (i.e., 276 tests) between the 24 lead SNPs prioritised by VariantSpark. BitEpi returned one significant interaction (*P*_FDR_ = 0.0105) after multiple testing correction: *rs*1970112 and *rs*74467064 (Supplementary Table 5, see file ‘Supplementary_Tables’). Consistent with an epistatic interaction, *rs*1970112 is in the intronic region of the long non-coding RNA (*CDKN2B-AS1*), while *rs*74467064 lies in the intronic region of *RBMS3,* a member of the RNA-binding motif (RBM) family which play roles in multiple biological activities including RNA stability and pre-mRNA splicing (29)

While intronic, *rs*1970112 has been associated to *CDKN2B-AS1* expression levels in aortic tissue in the Stockholm-Tartu Atherosclerosis Reverse Networks Engineering Task (STARNET) (30). The *CDKN2B-AS1* risk loci has been associated with CAD across multiple ancestries (6), independent of traditional CAD risk factors such as cholesterol and hypertension. On the other hand, *RBMS3* has not been directly linked to CAD but it has been marginally associated to carotid intima-media thickness in sub-Saharan Africans (31) and is significantly differentially expressed between patients with CAD and matched controls in STARNET across multiple tissues, including the aorta and liver.

Replication of the *rs*1970112 (*CDKN2B-AS1*) and *rs*74467064 (*RBMS3*) interaction in the independent All of Us dataset (AoU; *n* = 175,731) was tested using BitEpi only. However, this pair returned a non-significant result (*P* = 0.14), likely due to variance in allele frequencies and resulting difference in genotype combinations between datasets. We hence expanded the analysis to SNPs in moderate to high LD (0.2 < R^2^ < 0.5) to *rs*74467064 and *rs*1970112. Amongst the interactions tested, *rs*17665445-*rs*1970112 returned a significant BitEpi result (*P* = 0.048) but failed multiple testing correction (BitEpi *P*_FDR_ = 0.24) (Supplementary Table 6, see file ‘Supplementary_Tables’).

### AlphaFold3 predicts physical interaction between Rbms3 protein and *CDKN2B-AS1* RNA

To investigate the epistasis hypothesis further, we modelled the potential physical binding between *CDKN2B-AS1* and *RBMS3*. From the literature, *RBMS3*’s known roles in cancer progression is primarily through interactions with lncRNAs (32,33) and *RMBS3* has been implicated in epistatic interactions linked to other complex traits (34,35). Furthermore, *CDK2NB-AS1* has been shown to physically interact with *RBMS1* (36), a paralog of *RBMS3*.

Using AlphaFold3 (37), the physical interaction between *CDKN2B-AS1* lncRNA and Rmbs3 protein was modelled. Using a 3.5Å cutoff to define a contact, a threshold commonly used to capture atomic interactions such as hydrogen bonds and van der Waals contacts (38), there were 1,054 predicted contact points between the *CDKN2B-AS1* lncRNA and the Rbms3 protein (Supplementary Table 7, see file ‘Supplementary_Tables’). A particularly extensive interaction surface was identified and mapped to exon 19 of the *CDKN2B-AS1* transcript (NR003529.4). Interestingly, RBPmap (39) analysis significantly matched an experimentally defined *RBMS3* motif to this *CDKN2B-AS1* region (chr9:22120679-22120692; hg19) (Z = 2.28; P < 1.12 × 10^-2^; Supplementary Table 8, see file ‘Supplementary_Tables’). In the AlphaFold3 model, this region corresponds to a loop-like structure in *CDKN2B-AS1* suggesting a strong binding affinity to the Rbms3 protein (Figure 2).

**Figure 2.**
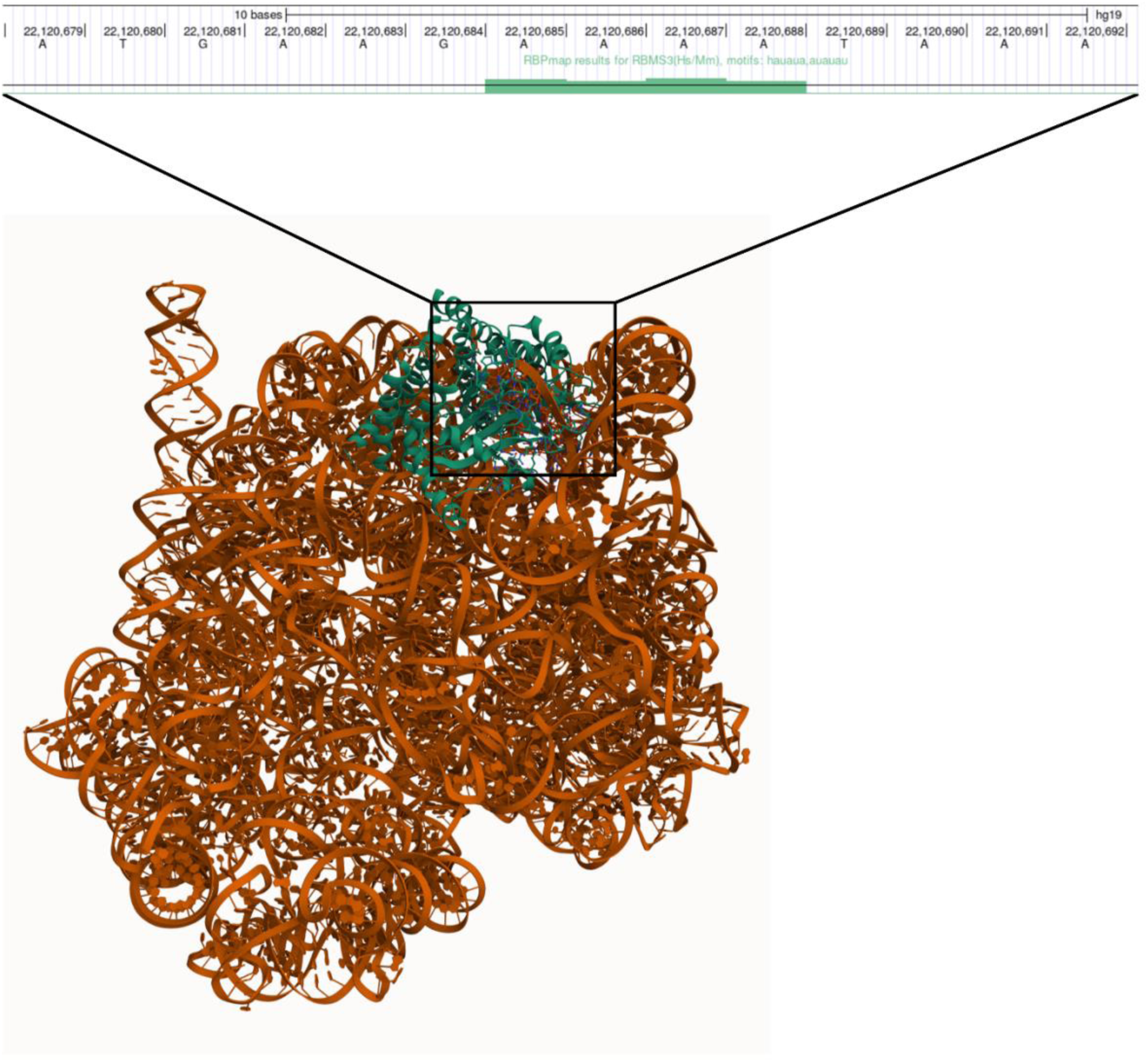
Screenshot of AlphaFold3 Prediction of CDKN2B-AS1 (NR003529.4) and Rbms3 (Q6XE24) Annotated with RBPmap Motif Analysis Result. AlphaFold3 was used to model CDKN2B-AS1 (coloured orange) and Rbms3 (coloured green) together. The loop-like structure in CDKN2B-AS1 mapped to a region in exon 19 of NR003529.4/CDKN2B-AS1. Motif analysis using RBPmap matched two experimentally defined RBMS3 motifs within this region in *CDKN2B-AS1* spanning chr9:22120679-22120692 (hg19/GRCh37).

As the SNPs identified in this epistatic interaction by VariantSpark-BitEpi (i.e., *rs*1970112 in *CDKN2B-AS1* and *rs*74467064 in *RMBS3*) are in intronic regions, they are not represented in the AlphaFold3 prediction as their roles are likely regulatory. In fact, their FORGEdb (40) scores are 7 and 6 (where 10 indicates highest likelihood of regulatory function), respectively, with established regulatory roles of *CDKN2B-AS1* and *RBMS3* in other polygenic traits, like cancer (41,42).

To evaluate the robustness of the AlphaFold3 prediction, we conducted additional modelling of Rbms1-*CDKN2B-AS1* as a positive control, previously shown to physically interact, and Actin-CDKN2B-AS1 as a negative control, experimentally demonstrated not to interact (36). AlphaFold3 predicted 93 Rbms1 protein residues in close spatial proximity to *CDKN2B-AS1*, compared with only 16 residues for Actin. This contrast can be visually discerned in Supplementary Figure 1 where the Rbms1 protein is entangled with *CDKN2B-AS1* while Actin is clearly separated from the RNA structure. Furthermore, the positive control (Rbms1) showed highly conserved RNA-binding residues which was not observed with the negative control (Supplementary Materials S1, see file ‘Supplementary_Materials’). Collectively, these findings suggest that AlphaFold3 does not generate spurious interaction when none exist, therefore supporting the biological plausibility of the predicted interaction between Rbms3 and *CDKN2B-AS1*.

## Discussion

In this study, we used a two-step approach to identify epistasis associated to CAD. We first applied VariantSpark to identify associations linked to CAD based on individual and epistatic effects before using BitEpi to exhaustively search for interactions within this narrowed search space. With this approach, we identified a putative epistatic interaction associated with CAD between two SNPs, *rs*1970112 (*CDKN2B-AS1*) and *rs*74467064 (*RBMS3*). While there likely are other interactions, we set a very stringent significance cut-off to only focus on a straightforward case for this paper.

*CDKN2B-AS1* (*ANRIL*) is a well-studied lncRNA, having been linked to various diseases including multiple cancer types (41,43), type 2 diabetes (44), and was one of the first CAD-risk loci that has also been consistently identified since (26,45). However, despite plausible involvement, its molecular mechanisms in CAD pathogenesis remain unclear due to the highly complex nature of that region. This includes multiple splicing events that generate various isoforms with opposing effects on atherosclerosis (46,47) and interactions with genes both within the 9p21.3 region (48) as well as in *trans* (49). While *RBMS3* has been primarily studied in cancer, where it exhibits both tumour-suppressive and oncogenic properties (42,50), it is also known to be involved in the Wnt/β-catenin signalling pathways (51) and angiogenesis (52), both of which have been implicated in CAD pathophysiology. *RBMS3* is also differentially expressed in CAD aortic tissue on STARNET. Both *CDKN2B-AS1* and *RBMS3* have been linked to breast cancer, which itself has a genetic causal relationship with CAD (53).

Furthermore, AlphaFold3 predicted an extensive interaction surface between *RBMS3* and *CDKN2B-AS1*. This interaction surface on *CDKN2B-AS1* mapped to the region that was significantly matched to an experimentally defined *RBMS3* motif by RBPmap. Moreover, *RBMS3*’s paralog, *RBMS1*, has been experimentally shown to bind to *CDKN2B-AS1* (36). At the cellular level, both the Rbms3 protein and *CDKN2B-AS1* transcripts have been detected in the cytoplasm (54,55), further suggesting the potential of a physical interaction. Collectively, these findings provide compelling evidence that *RBMS3* is a novel contributor to the genetic architecture of CAD but hasn’t been associated in previous GWAS studies as its effect is likely to be in epistasis with *CDKN2B-AS1*.

While replication of genetic associations across two or more independent datasets is expected in GWAS, for epistasis analysis, it is highly unlikely that the same combination of SNPs would be associated in the same model across two datasets due to variation in allele frequency and linkage disequilibrium (LD) patterns (56), and gene-based replication should be considered. This variation in MAF is something we observed in our study where *rs*74467064 had a MAF = 5.6% in the UKB cohort (i.e., considered a common variant) but a MAF = 4.2% in the AoU cohort (i.e., considered a low-frequency variant and may be filtered out). We, hence, expanded our analysis to evaluate combinations of SNPs in LD to *rs*74467064 and identified the pairwise interaction between *rs*17665445 (in LD with *rs*74467064, R^2^ = 0.3645, *p* < 0.0001) and *rs*1970112. However, this approach required multiple testing correction and *rs*17665445-*rs*1970112 did not stand up to that scrutiny (BitEpi *P*_FDR_ = 0.24). This highlights the importance of reducing the epistasis search space to minimise the multiple testing burden for effective epistasis detection as discussed in a review by Balvert et al (57). A better approach would have been to run the VariantSpark and BitEpi pipeline on the AoU dataset to pinpoint the SNPs in *RBMS3* and *CDKN2B-AS1* that are working together in epistasis specific to the AoU dataset. However, we were unable to do so due to limited access to AoU, which forms a limitation of this study.

Another interesting discovery is VariantSpark’s increased power in identifying associations, aligning with our previous findings in Alzheimer’s disease (16). Specifically, on the TM cohort, VariantSpark was able to identify both *LPA* and *CDKN2B-AS1* risk loci while PLINK only identified *CDKN2B-AS1*. This is even more pronounced on the larger UKB dataset (UKB cohort, *n* = 51,107) with a well-defined phenotype. Here, VariantSpark identified CAD risk loci that were previously only detected in substantially larger cohorts, including *PMAIP1-MC4R* identified in 2015 with 184,505 samples (58), *AAK1* in 2022 with 1,559,665 samples (5), and *WWP2* in 2022 with 773,238 samples (6). None of these were identified by PLINK’s logistic regression on the UKB cohort. This increase in power is likely due to VariantSpark’s ability to detect both individual and interactive effects as well as capture non-linear relationships.

## Conclusions

In conclusion, we have demonstrated the effectiveness of a two-stepped approach for epistasis detection in CAD, combining ML-based genome-wide loci prioritisation with exhaustive search for interactions. The first step leverages the higher sensitivity of VariantSpark over logistic regression to identify genetic variants associated with CAD, reducing the cohort sizes needed for epistasis detection. In combination with BitEpi, we uncovered *RBMS3* as a novel CAD-risk gene, working in epistasis with *CDKN2B-AS1*, a well-established but mechanistically unclear CAD risk locus. We show strong *in silico* evidence in support of this epistatic interaction, including AlphaFold3 predictions of physical interaction between *Rbms3* protein and a *CDK2B-AS1* isoform, warranting further experimental validation. These findings illustrate the potential of our approach, and with expanding cohort sizes will enable the discovery of additional epistasis, addressing the missing heritability of CAD and other complex traits.

## Methods

### Datasets and Cohort Selection

#### GWAS & Epistasis Discovery using UK Biobank

Detailed information regarding the UK Biobank (UKBB) study are as described by Bycroft et al.(19), however, a summary is provided here. Between 2006 and 2010, the UK Biobank recruited approximately 500,000 participants aged between 40 and 69 from England, Wales, and Scotland. A variety of phenotypic information and biological samples were collected and analysed at recruitment. In addition, participants’ electronic health records, which included inpatient International Classification of Disease (ICD) codes and OPCS Classification of Interventions and Procedures (OPCS-4) codes, were linked and incorporated.

Coronary artery disease patients were identified using the following ICD-9 and ICD-10 codes: 410, 411, 413, 414; I20-25, Z951, Z955, and the following OPCS-4 codes: K40-46, K49, K50, K75.

The control group were non-smoking participants with a calculated BMI < 30 and without diagnoses of other cardiovascular diseases and known comorbidities, including epilepsy and heart arrhythmias.

Samples that passed the following quality control criteria were kept: (i) not carriers of full or mosaic sex chromosome aneuploidies, (ii) matched reported and genetic sex, (iii) without genetic kinship to other UKBB participants, (iv) missing call rates > 98%, (v) within +/-3 standard deviation of calculated heterozygosity rate mean, (vi) identified as white British with similar genetic ancestry based on principal component analysis of UKBB genotypes.

The resulting number of cases were 25,107 with 157,665 available controls. To keep the ratio of case to controls 1:1, 26,000 controls were chosen at random.

#### GWAS Validation using TOPMed

The TOPMed program sits under the broader precision medicine initiative of the National Institutes of Health (NIH) aimed to tailor disease treatments to an individual’s unique genomics and environment. Detailed information regarding the TOPMed study is as described by (20). In brief, high-coverage whole-genome sequencing (WGS) was completed on 53,831 TOPMed samples across 32 participating studies with deep phenotyping to understand risk factors for complex disorders, including heart disease. As each participating study collected phenotypic data such as physical measurements, clinical chemistry, and clinical registries individually, the TOPMed Data Coordinating Center (DCC) has created harmonized phenotypes for combined analyses.

Access to five TOPMed datasets and their parent study was approved (Project #29236) and downloaded through dbGaP, including the Atherosclerosis Risk in Communities (ARIC) study, the Coronary Artery Risk Development in Young Adults (CARDIA) study, the Cardiovascular Health Study (CHS), the Framingham Heart Study (FHS), and the Women’s Health Initiative (WHI) study.

Coronary artery disease patients were identified using the following harmonised datasets: atherosclerosis events incidents and prior except for peripheral arterial disease indicators. Where available, ICD-9 and ICD-10 codes were also extracted from individual studies where CAD patients were identified using the same codes as with the UK Biobank.

The control group were non-smoking participants with BMIs < 30 and without indications of other cardiovascular diseases including hypertension and hyperlipidaemia.

Samples were passed through the following quality control measures: (i) no sex aneuploidy, (ii) matched reported and genetic sex, (iii) unrelated to the second degree to other participants, (iv) missing call rates > 98%, (v) within ±3 standard deviations of calculated heterozygosity rates, (vi) clustered tightly with the 1000 Genomes Project European population on a principal component plot (PC1 vs PC2).

The final number of samples were 11,326 with 5,509 cases of CAD.

#### Epistasis Validation using All of Us

The All of Us Research Program (All of Us) is a longitudinal study aimed at enrolling a diverse cohort of at least one million individuals across the United States to accelerate biomedical research and enhance human health(59). Since its launch in May 2018, All of Us has enrolled over 633,000 participants aged 18 and older from more than 340 recruitment sites nationwide, contributing to the Curated Data Repository (CDR, Control Tier Dataset v8) released in 2025. The program integrates multiple data modalities, including electronic health records, survey responses (participant-provided information), physical measurements, biospecimens, and data from digital health technologies such as wearables.

Coronary artery disease patients were identified using the following ICD-10 codes: I20-25, Z951 and Z955. The control group were non-smoking participants with a calculated BMI < 30 and without diagnoses of other cardiovascular diseases and known comorbidities, such as hypertensive disorder and diabetes.

Samples that passed the following quality control criteria were kept: (i) not carriers of full or mosaic sex chromosome aneuploidies, (ii) matched self-reported and genetic sex, (iii) without genetic kinship to other All of Us participants, (iv) missing call rates > 98% and (v) (v) Allele balance (AB) ≥ 0.2 for heterozygotes.

The resulting number of cases was 41,525 with 134,206 available controls.

### Genotyping and Quality Control

#### GWAS & Epistasis Discovery using UK Biobank

UKBB samples were genotyped using either the Affymetrix UK BiLEVE Axiom or the Affymetrix UK Biobank Axiom array and mapped to the GRCh37 reference. 806,466 directly genotyped DNA sequence variants passed variant quality control. The UKBB team then performed imputation from a combined 1000 Genomes and UK10K reference panel using SHAPEIT3 and IMPUTE3.

The following variant quality control filters were applied to the imputed genotypes: (i) imputation quality < 0.5, (ii) Hardy-Weinberg equilibrium *P* < 1 x 10^-6^, (iii) MAF < 0.01, (iv) call rate > 98%. Only bi-allelic single nucleotide polymorphisms (SNPs) were kept resulting in a total of 4,253,140 genetic variants.

#### GWAS Validation using TOPMed

Standardised laboratory methods, a single pipeline of mapping and processing of WGS data to the GRCh38 reference, and joint variant calling and genotyping across all participating studies, year, and sequencing centres were performed to minimise batch effects. Stringent quality filters were used to ensure that batch effects were minimised and that genotype calls were of high quality. On average, 99.65% of the reference genome was covered to a mean read depth of 38.2x resulting in 3,748,599 variants per individual.

The freeze 8 genotype data of the five TOPMed datasets accessed were downloaded individually and merged into a single VCF. The following variant quality control filters were then applied to the merged VCF: (i) Hardy-Weinberg equilibrium *P* < 1 x 10^-6^, (ii) MAF < 0.01, (iii) call rate > 98%, (iv) bi-allelic single nucleotide polymorphisms (SNPs). As the TOPMed cohort was used to validate the UK Biobank findings, the genotype calls were mapped to the GRCh37 reference using liftOver (60) and subset to include SNPs also found in the UK Biobank genotype data resulting in 3,996,295 genetic variants.

#### Epistasis Validation using All of Us

The All of Us dataset contains short-read whole genome sequencing (srWGS) data for 414,830 participants, including more than 1.2 billion single nucleotide variants (SNVs) and indels, with multiallelic sites split into separate records. The dataset includes three callsets, and the Allele Count/Allele Frequency (ACAF) threshold callset was selected for this analysis. This callset includes variants with a population-specific allele frequency (AF) greater than 1% or a population-specific allele count (AC) greater than 100 in any of the computed ancestry subpopulations. The prioritised pairwise interactions from the BitEpi analysis on the UK Biobank cohort was filtered and then used to construct contingency tables (3 x 3 for each genotype combination) for CAD cases and controls.

### GWAS using Logistic Regression and VariantSpark

#### Discovery using UK Biobank

A logistic regression model between the 25,107 CAD cases and 26,000 controls and the dosage of 4,253,140 genetic variants (nV) was performed. Age, sex, genotype array, BMI, and the first 10 principal components were included as covariates. The widely accepted genome-wide significance threshold (i.e., 5 × 10^-8^) was used to correct for multiple-testing.

For the VariantSpark implementation, genotype hard calls were used instead of dosages. This resulted in some imputed genotype dosages to be coded as missing hard call genotypes due to a hard call threshold of 0.1 (i.e., 0.5 × ∑_*i*_ |*x*_*i*_ − ⌊*x*_*i*_⌋| ≯ 0.1). The resultant missing hard call genotypes were imputed using VariantSpark’s inbuilt ‘mode’ imputation which replaces missing genotypes with the most frequent call of non-missing genotypes. Following hyperparameter optimisation (Supplementary Material S2, see file ‘Supplementary_Materials’), the following parameters for the final VariantSpark model were used: nTree = 15,000, mTry = 0.1 x nV, maximum depth = 13, minimum node size = 10,000.

*P*-values were calculated using the RFlocalfdr approach(61) and a false discovery rate of 5% was used to denote significance. The significant SNPs were then clumped using PLINK’s --clump function with the following settings: --clump-kb 5000 –-clump-r2 0.1 and the 1000 Genomes Project European cohort. A locus is therefore defined as a 1MB region about the most significant SNP within the haplotype.

#### Validation using TOPMed

A logistic regression model between the 5,509 cases and 5,817 controls and the genotype hard calls of the 3,996,295 SNPs that overlapped with the UK Biobank was performed. Age, sex, BMI, and the first 10 principal components were included as covariates. The widely accepted genome-wide significance threshold (i.e., 5 × 10^-8^) was used to correct for multiple-testing.

The same dataset was used in VariantSpark and as none of the SNPs were missing data, no imputation was required unlike with the UK Biobank cohort. Following hyperparameter optimisation (Supplementary Material S3, see file ‘Supplementary_Materials’), the following parameters were used: nTree = 15,000, mTry = 0.1 x nV, maximum depth = 13, minimum node size = 1,000.

Similarly, *P*-values were calculated using the method outlined in Dunne et al. (2022)(61) and a false discovery rate of 5% was used to denote significance. The significant SNPs were then clumped using PLINK’s –-clump function with the following settings: --clump-kb 5000 -- clump-r2 0.1 and the 1000 Genomes Project European cohort, defining a locus as the 1MB region about the most significant SNP within the haplotype.

### Epistasis Analysis using BitEpi

#### Discovery using UK Biobank

After clumping the 115 significant VariantSpark SNPs, the resulting 24 independent SNPs were exhaustively analysed with BitEpi(18) for pairwise interactions (i.e., 276 tests). Briefly, the β-value, an entropy metric based on the concept of set-purity, quantifies the marginal and interaction effect of the pair and is used to compute the α-value, the interaction effect size for each interaction. In this study, all component α-values of the interaction are subtracted, rather than the maximum combined association power of any subset only as in Bayat et al., 2021(18) to calculate a more accurate measure of the interaction effect size. Individual genotype numbers in contingency tables were also altered to remove the correlation between SNPs while preserving individual allele frequencies and case-control ratios. A permutation approach where the case-control phenotype was randomly permuted 1,000,000 times and the α-value calculated for each permutation was used to estimate the *P* value. The *P* value is calculated as the number of permutations where the permuted α-value is larger than the true the α-value divided by the total number of permutations (i.e., 1e^6^).

#### Validation using All of Us

BitEpi was run on the contingency tables of the following pairs which were generated from the All of Us dataset: (i) *rs*74467064-*rs*1970112, (ii) *rs*9864192-*rs*1970112, (iii) *rs*17665445-*rs*1970112, (iv) *rs*9310894-rs1970112, (v) *rs*74467064-*rs*1537370. All values (β, α, and *P*) were calculated as described above with 1e^6^ permutations for *P*-value estimations. Additionally, the chi2_contingency function from the Python Scipy library was used to compute the chi-square statistic and p-value for the chi-square test of independence.

### Downstream Analysis

#### Variant to Causal Gene

Significantly associated SNPs were functionally annotated using the two mapping approaches: positional and molecular phenotype quantitative trait loci datasets. Firstly, based on chromosomal position, the variants were mapped to dbSNP v. 150 for rsIDs, RefSeq genes, and functional predictions using ANNOVAR (62) (version April 2021). The Stockholm-Tartu Atherosclerosis Reverse Networks Engineering Task (STARNET; http://starnet.mssm.edu/) was used to determine the expression quantitative trait loci information of associated SNPs.

#### Structure Predictions and Motif Analysis

AlphaFold3 was used to predict three interactions with six seeds per interaction. The best model (seed) for each interaction was chosen through AlphaFold3’s ranking score which reflects the overall quality and physical plausibility of the predicted interactions. The three interactions were: 1) *CDKN2B-AS1* NR003529.4 transcript sequence in FASTA format and Rbms3 protein sequence UniProt (UniProt ID = Q6XE24), 2) *CDKN2B-AS1* NR003529.4 transcript sequence in FASTA format and Rbms1 protein (UniProt ID = P29558), and 3) *CDKN2B-AS1* NR003529.4 transcript sequence in FASTA format and Actin protein (UniProt ID = P60709). The resulting .cif files was input into Mol* viewer(63) to visualise, identify contact points, find conversed RNA-binding protein residues, and to identify the regions mapping to structural points of interest. The genomic coordinates (GRch38/hg38) chr9:22120503-22120712:+ was used as the query coordinates for RBPmap(39) and RBMS3 from the RBPmap motifs database for motif selection. Default settings for all other options were kept. RBPmap results were visualised in the UCSC Genome Browser through the inbuilt option. The SNPs rs74467064 and rs1970112 was queried on FORGEdb’s web server to gain their FORGE2 score.

## Data Availability

Access to the UK Biobank dataset is upon application and with permission of UKB Research Ethics Committee. Access to the TOPMed dataset is through dbGaP. This study was conducted under the approved project 29236. Access to the All of Us cohort is upon application. This study was conducted through Cedars-Sinai Medical Center.

http://www.ukbiobank.ac.uk/using-the-resource

https://topmed.nhlbi.nih.gov/topmed-data-access-scientific-community

https://www.researchallofus.org/register/

## Data Availability

Access to the UK Biobank dataset is upon application as described at http://www.ukbiobank.ac.uk/using-the-resource and with permission of UKB’s Research Ethics Committee. Access to the TOPMed dataset is through dbGaP and is upon application as described at https://topmed.nhlbi.nih.gov/topmed-data-access-scientific-community. This study was conducted under the approved project #29236. Access to the All of Us cohort is upon application as described at https://www.researchallofus.org/register/. This study was conducted through Cedars-Sinai Medical Center.

## Code Availability

VariantSpark (v0.5.3) is freely available on GitHub: https://github.com/aehrc/variantspark and on AWS and Azure marketplace. An example Jupyter notebook is included in the GitHub repositories for reference and is reflective of the Jupyter notebook used for this study. Operating system: Linux. Programming language: Python, HAIL, Bash. Other requirements: Scala, Apache Spark, htsjdk, args4j, Joda-Time, fastutil, scala-csv. License: CSIRO Open Source Software Licence v1.0.

BitEpi is available on GitHub https://github.com/aehrc/bitepi respectively. Operating system: Platform independent. Programming language: Python. Licence: CSIRO Open Source Software Licence v1.0

## Author Contribution

J.W.V, D.C.B., & N.A.T. conceived of this study. B.H., P.S., R. R., & Y.J. developed VariantSpark and BitEpi. L.M.F.S ran all analyses with contributions from M.O.B. for the TOPMed dataset, M.V., P.J.F, Z.W., & J.H.M for the All of Us analysis, and A.H.K. & M.K. for AlphaFold3 analysis. L.M.F.S wrote the manuscript with contributions from A.P.. All authors reviewed and approved the final draft of the report.

## Acknowledgements

This study was conducted using the UK Biobank Resource under Application Number 27483.

## Additional files

Description of data

1. File name: Supplementary_Materials.docx Title of data: Supplementary Materials Description of data: Additional material in support of the findings in primary manuscript, including hyperparameter optimisation for VariantSpark and AlphaFold3 analysis.
2. File Name: Supplementary_Tables.xlsx Title of data: Supplementary Tables Description of data: Additional material in support of the findings in the primary manuscript, including associated variants from VariantSpark and Logistic Regression analysis, BitEpi results and AlphaFold3 results.

