## Supplementary figures and images for "Novel Epistatic Interaction Between RBMS3 and CDKN2B-AS1 in Coronary Artery Disease Risk Identified by Machine Learning Tool VariantSpark"

### Supplementary Figure 1

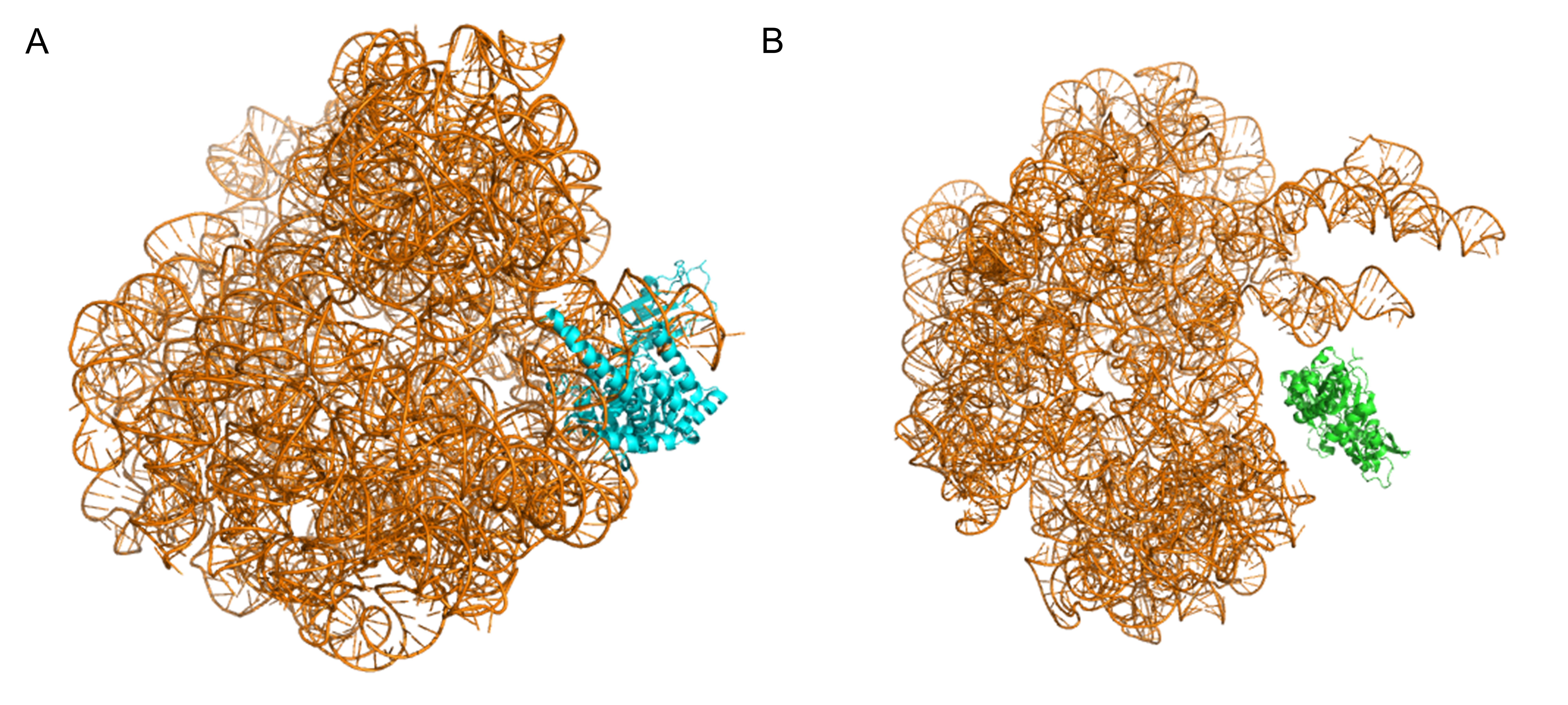
