## Supplementary Information for "Novel Epistatic Interaction Between RBMS3 and CDKN2B-AS1 in Coronary Artery Disease Risk Identified by Machine Learning Tool VariantSpark"

S1 **Additional AlphaFold Analysis**

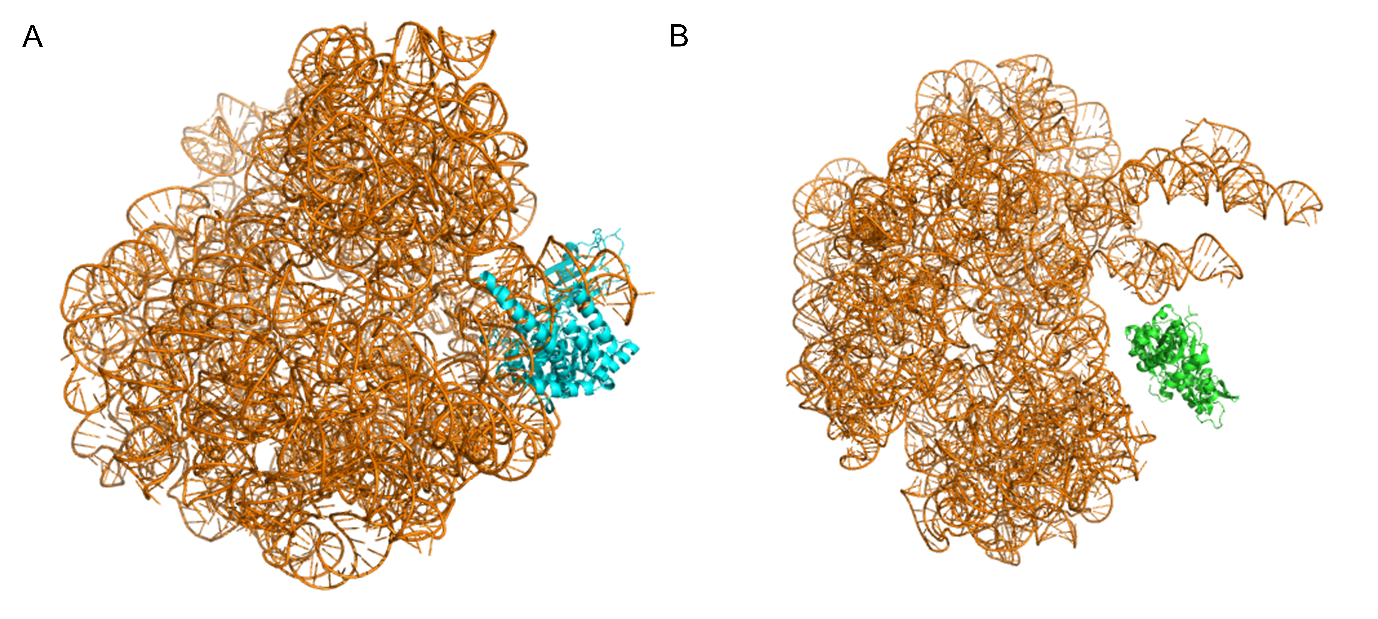

SFigure 1. Screenshots of AlphaFold3 Prediction of CDKN2B-AS1 (NR003529.4) with (A) Rbms1 and (B) Actin. AlphaFold 3 was used to model (A) CDKN2B-AS1 (coloured orange) and Rbms1 (coloured blue) together and (B) CDKN2B-AS1 (coloured orange) and Actin (coloured green). AlphaFold3 predicted close physical contact between CDKN2B-AS1 and Rbms1 but none between CDKN2B-AS1 and Actin which is as expected as experimental evidence has shown a physical interaction between CDKN2B-AS1 and Rbms1 but not with Actin.

Six AlphaFold3 models were used for conserved RNA-binding coverage analysis for each of the following interactions: 1) Rbms1 & *CDKN2B-AS1* and 2) Actin & *CDKN2B-AS1*. There were 15 highly conserved RNA-binding protein residues between Rbms1 and *CDKN2B-AS1* (STable 1) which were consistently predicted by all 6 models but 0 between Actin and *CDKN2B-AS1*.

STable 1. List of 15 Highly Conserved RNA-Binding Protein Residues. These were based on six AlphaFold3 models of Rbms1 and *CDKN2B-AS1*.

| Protein Location | Amino Acid | Number of Models (out of 6) | Percentage of Models |
| --- | --- | --- | --- |
| 134 | LYS | 6 | 100 |
| 135 | GLN | 5 | 83 |
| 136 | GLN | 6 | 100 |
| 144 | TYR | 6 | 100 |
| 171 | ARG | 6 | 100 |
| 173 | LEU | 6 | 100 |
| 174 | ARG | 6 | 100 |
| 175 | ASP | 5 | 83 |
| 181 | ARG | 6 | 100 |
| 183 | VAL | 5 | 83 |
| 185 | PHE | 6 | 100 |
| 225 | ASP | 6 | 100 |
| 226 | GLY | 5 | 83 |
| 227 | GLY | 5 | 83 |
| 231 | ARG | 5 | 83 |

S2 **Hyperparameter Optimisation for VariantSpark with UK Biobank Data**

Hyperparameter optimisation was conducted in R/v4.2.1^1^ with the R library **ranger**^2^ using the QC’d UK Biobank (UKB) CAD cohort (number of samples = 51,107 and number of genetic variants = 4,253,140). Batches of random forests (RFs) with various hyperparameter settings described below were built. The advantage of building batches of RFs rather than single RF models is that bootstrapping of the model metric out-of-bag error (OOB-ER) can be performed and the variability/stability of the metric can be evaluated.

The following three key hyperparameters were optimised:

1. nTree: Number of trees in the forest
2. minNS: Minimum number of observations in a node to be processed.
3. mTry: *Fraction* of total number of variants evaluated at each node of a tree

The OOB-ER, an unbiased estimate of the generalised error of the RF model, was used to optimise the hyperparameters. The bootstrapping of OOB-ER was also performed with 1,000 folds. Note that during bootstrapping, when nTree equal the total number of trees available, all available batches of RFs are used for every bootstrap sample resulting in no variability and SEM = 0. Furthermore, as RFs are relatively robust against hyperparameters, the variation in OOB-ER between RFs is minute (range = 0.466 - 0.471).

The following sections (S1.1, S1.2 S1.3) will show that using OOB-ER, these following values are optimised for each hyperparameter:

- nTree = 10,000 – 15,000
- minNS = 10,000
- mTry = 0.1*nV
  S2.1 **nTree**

It is accepted that the larger the nTree value to better, but it is also known, that with larger nTree comes a computational cost that results in diminishing returns. Furthermore, compared to building prediction models, variable identification tend to require larger forests^3^. This requirement of large forests is especially true for the RFlocalfdr method used in the analysis to calculate *p*-values^4^.

A total of 250 batches of RFs were built with the following parameters:

- nTree = 100
- mTry = 0.1*nV (425,314)
- minNS = 10,000

Therefore, when the batches were joined, the cumulative number of trees evaluated is 25,000.

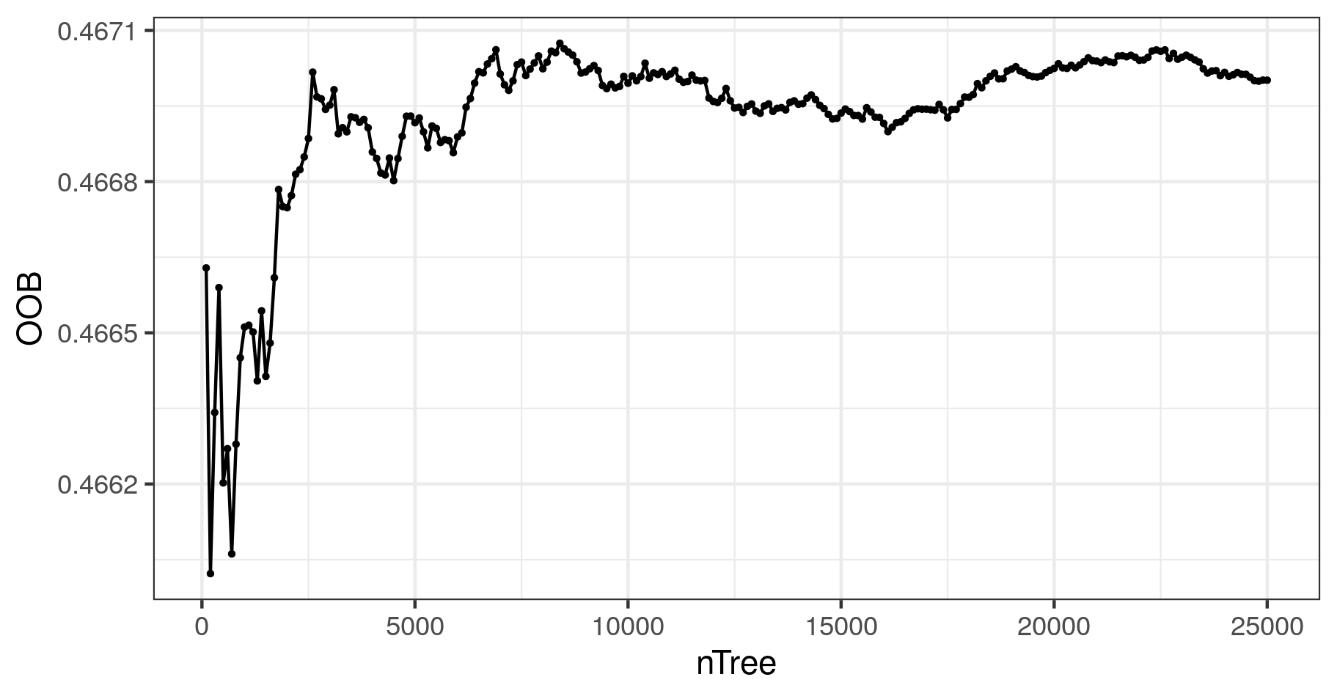

SFigure 2. nTree vs OOB-ER with UKB data. As expected, OOB-ER is very variable when nTree is small (i.e., < 2,500) but starts to stabilise when nTree ≈ 10,000. Note that the differences in OOB-ER (*y*-axis) are slight, within 0.003, due to the robustness of random forests against the effect of hyperparameters. The figure suggests that increasing nTree over 10,000-15,000 would result in more computational load for limited gains in OOB-ER.

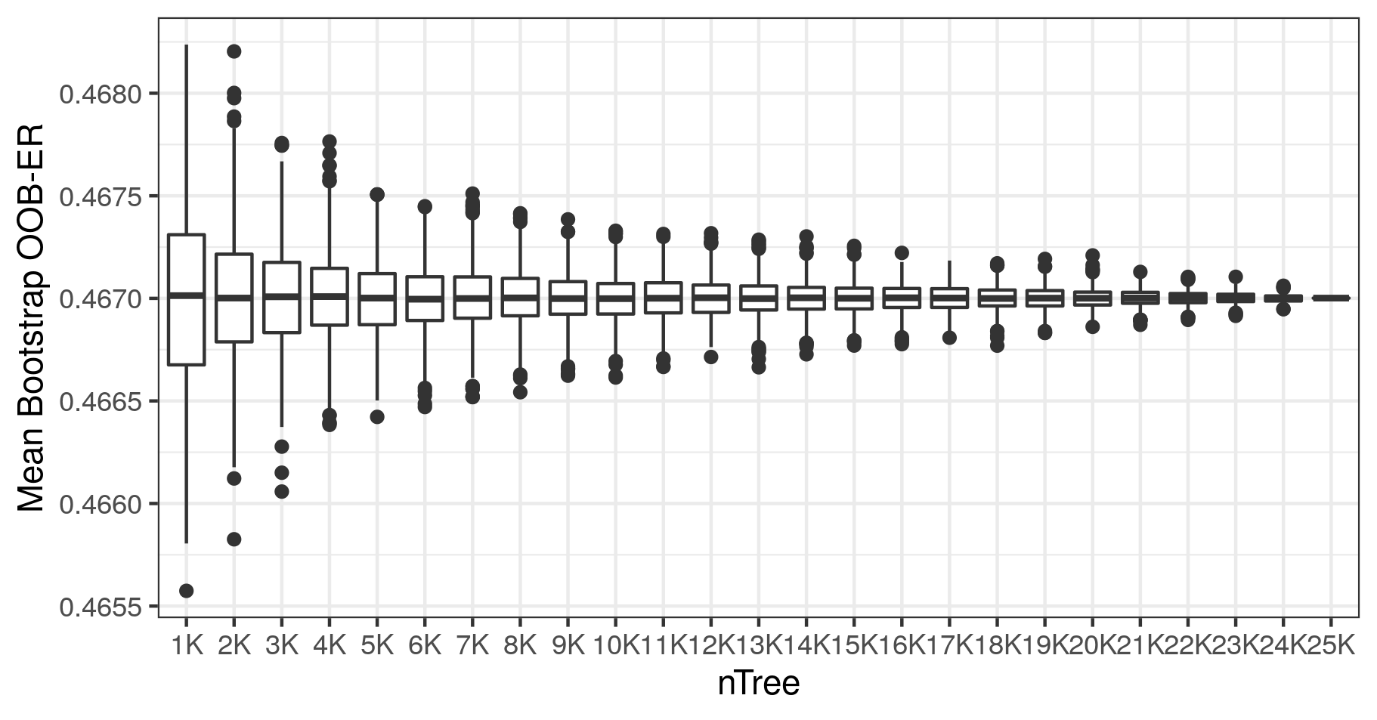

SFigure 3. nTree vs Mean Bootstrap OOB-ER with UKB data. In concordance with SFigure 1, the variability of OOB-ER is large when nTree is smaller (e.g., < 3,000) but starts to become comparable as it gets larger. The distribution of OOB-ER across bootstraps are similar when nTree ~ 9,000 – 18,000) suggesting limited gains in building more trees pass nTree ~ 9,000.

As expected, OOB-ER decreases with increasing nTree. From SFigure 1, OOB-ER stabilises when nTree ≈ 10,000. Furthermore, with increasing nTrees we have more confidence in the OOB-ER as seen by the decreasing range and number of outliers in the boxplots (SFigure 2A). The elbow of SFigure 2B occurs at about nTree = 8,000, suggesting that this is where the decrease in SEM does not justify the cost of building more nTrees. It is worth noting that differences in OOB-ER is small, as evident in the *y*-axis of SFigure 1 and SFigure 2.

S2.2 **Minimum Node Size (minNS)**

minNS is a parameter that controls tree size and is usually set to a default value of 1 (for classification trees) as unpruned trees have increase variance^3^. On the other hand, limiting tree size can speed up computation and it has been shown that when the data is noisy (e.g., genotype data), performance may improve with large minNS thereby limiting the number of superfluous splits and resulting in smaller trees with only the important predictors^5^. Tree depth is another hyperparameter that controls the depth/size of tree, but simulations and theoretical analysis of median forest showed that tuning either minNS or tree depth was sufficient for similar performance^6^.

Again, a total of 250 batches of RFs were built with the following parameters:

- nTree = 100
- mTry = 0.1*nV (425,314)
- minNS = 5,000, 10,000, 20,000

Therefore, when the batches were joined, the cumulative number of trees evaluated is 25,000.

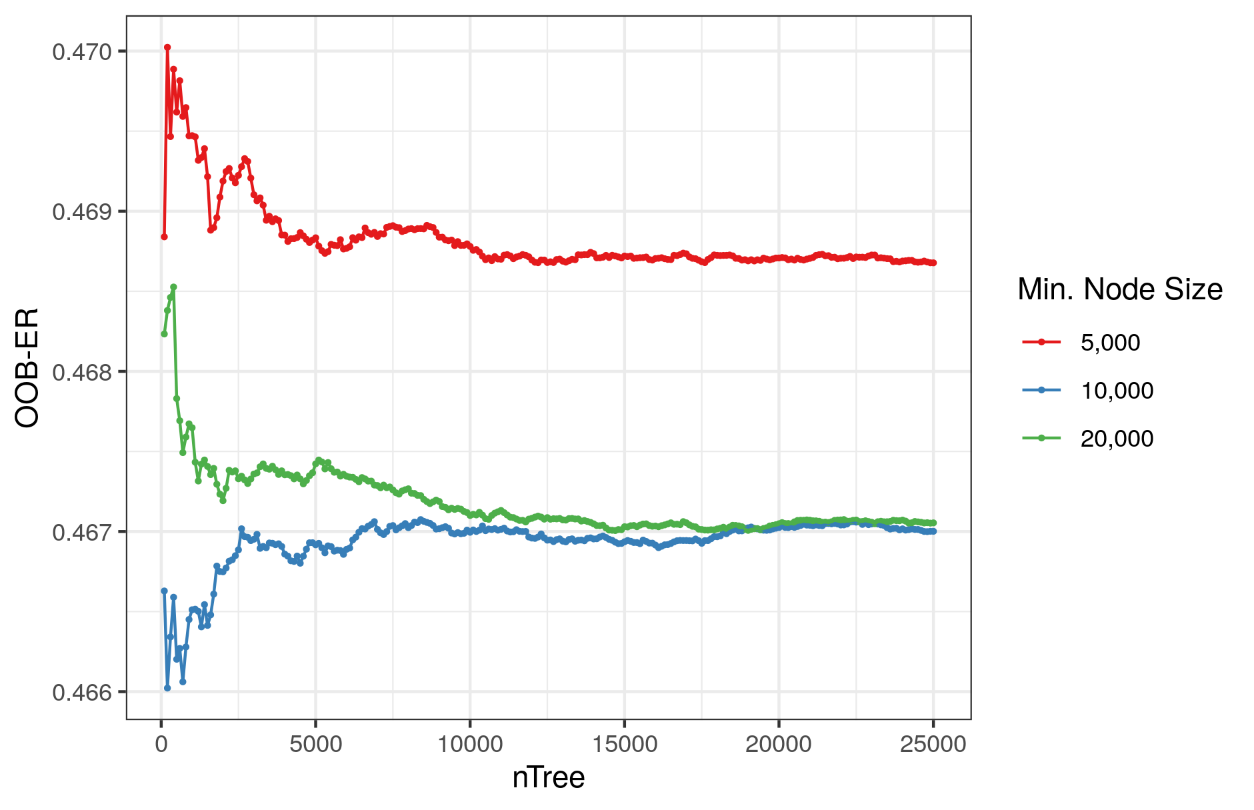

SFigure 4. OOB-ER vs nTree for various minNS with UKB data. As expected, OOB-ER stabilises for all minNS when nTree gets relatively large (e.g., ~5,000). On the other hand, OOB-ER decreases as minNS increases although not sequentially depending on nTree (e.g., when nTree = 5,000, minNS = 20,000 had a larger OOB-ER than minNS = 10,000 but smaller OOB-ER than minNS = 5,000). However, when nTree ~ 10,000 and greater, OOB-ER is comparable when minNS = 10,000 and 20,000.

From SFigure 3, there is a clear decrease in OOB-ER between minNS of 5,000 and the larger minNS (i.e., 10,000 and 20,000). However, between minNS of 10,000 and 20,000 the difference in OOB-ER is not as stark, especially when nTree gets greater than 10,000. Interestingly, when nTree is less then 10,000, RFs with 10,000 minNS resulted in smaller OOB-ER then RFs with 20,000 minNS suggesting that it can be just as detrimental to build RFs too shallow as when they are built too deep.

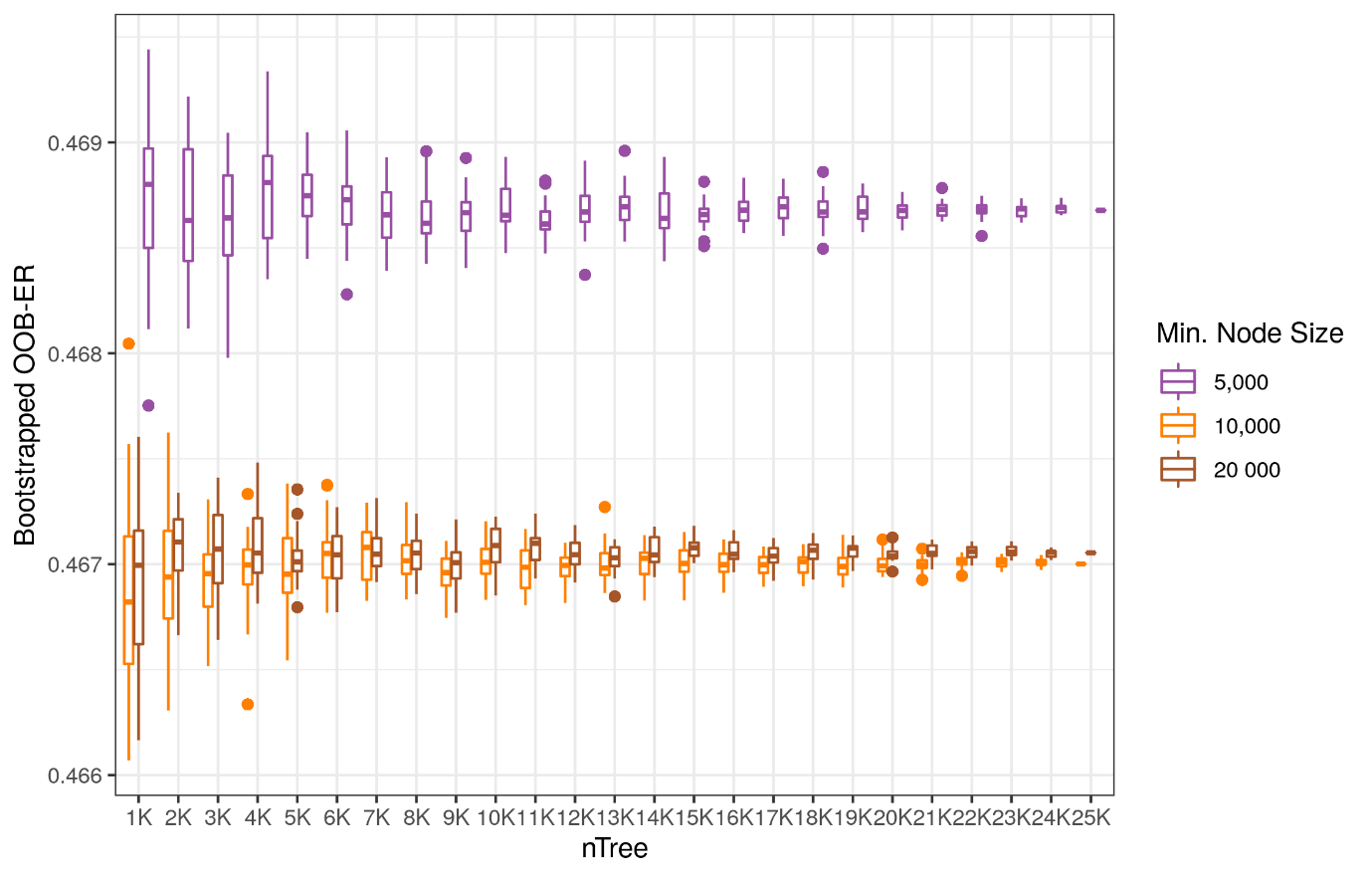

SFigure 5. Bootstrapped OOB-ER vs nTree for various minNS with UKB data. Like SFigure 2, variability in OOB-ER decreases as nTree increases for all minNS trialled. There is clear separation of OOB-ER for all nTrees between minNS = 5,000 and minNS 10,000 or 20,000. However, there seems to be no difference in OOB-ER for all nTree when minNS = 10,000 and 20,000, as indicated by the overlapping boxplots.

Although SFigure 3 suggests that using minNS of 10,000 when nTree < 10,000 will result in lower OOB-ER compared to the other tested minNS values, SFigure 4 shows that a minNS of 20,000 has a similar range of OOB-ER for nTree. This suggests that there wouldn’t be much difference in OOB-ER between minNS of 10,000 and 20,000 and that the decision between the two minNS options can be based on other variables including computational time and cost.

S2.3 **mTry**

mTry has the greatest potential impact on the complexity of the RF model with smaller mTrys producing less correlated trees, thereby reducing the overall variance of the prediction but at the cost of increased bias^3^. Although an mTry value of $\sqrt{nVariables}$ was first advised as a good starting point, when there are many weak predictors, this value would need to be increased^7^. This has been shown to be true with high-dimensional genotype data^8^ where an mTry of 0.1*nSNP resulted in a substantial improvement in accuracy compared to the recommended $\sqrt{nVariables}$.

Unlike previous sections, a total of 240 batches of RFs were built instead with the following parameters:

- nTree = 100
- mTry = 0.1*nV (425,314), 0.2*nV(850,628), 0.5*nV (2,126,570)
- minNS = 5,000, 10,000, 20,000

Therefore, when the batches were joined, the cumulative number of trees evaluated is 24,000.

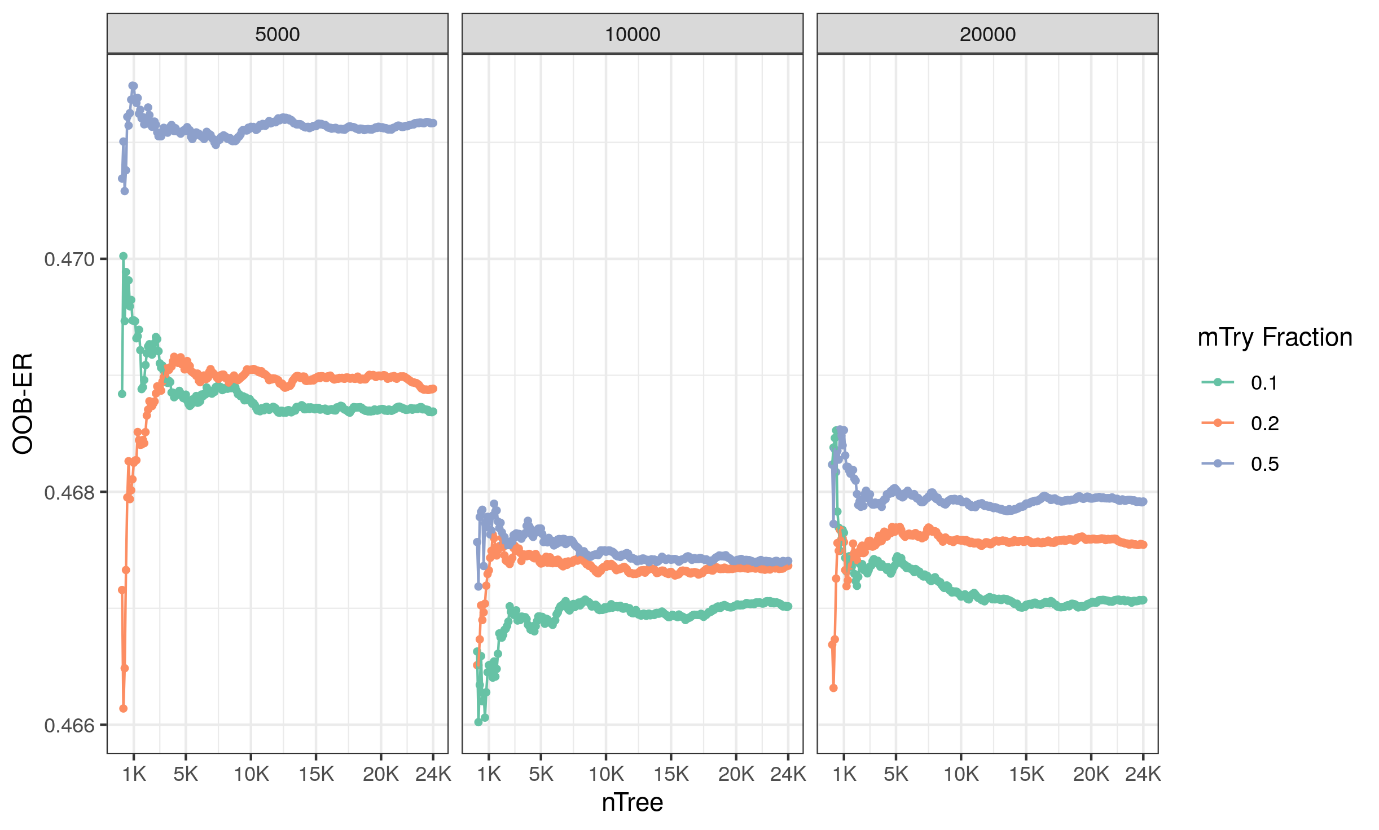

SFigure 6. nTree vs OOB-ER for various mTry values faceted by minNS with UKB data. Unsurprisingly, the plot above suggests an interplay between the nTree, mTry, and minNS hyperparameters. RFs with minNS = 5,000 resulted in higher OOB-ERs for all mTry when nTree was sufficiently large (e.g., 5,000). As suggested in SFigure 4, there seems to be overlap in OOB-ER between minNS = 10,000 and 20,000, however, this depends on mTry fraction. For example, RFs with smaller mTry (i.e., 0.1) and larger minNS (i.e., 20,000) have comparable OOB-ER to RFs with larger mTry (i.e., 0.2 and 0.5) but smaller minNS (i.e., 10,000). Ultimately. The RF with a minNS = 10,000 and mTry = 0.1 resulted in the lowest OOB-ER, although when nTree > 15,000 the RF with minNS = 20,000 and mTry = 0.1 also resulted in comparable OOB-ER.

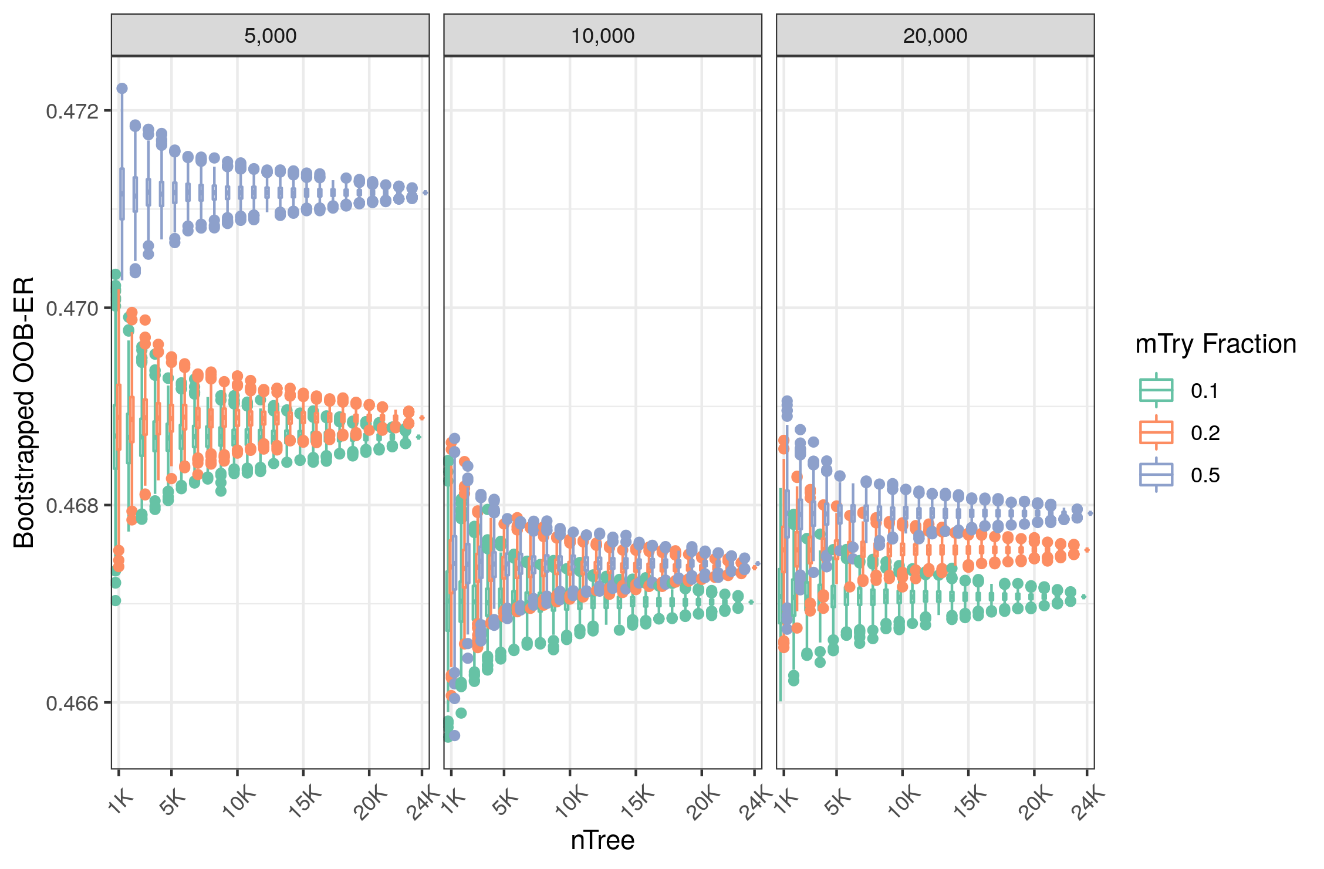

SFigure 7. nTree vs Bootstrap OOB-ER for various mTry faceted by minNS with UKB data. There is high OOB-ER overlap between mTry = 0.1 and 0.2 for all minNS and nTree. This is also true for mTry = 0.5 when minNS = 10,000 and 20,000, although there is some separation of OOB-ER between mTry when nTree > 20,000. RFs with mTry = 0.1 and minNS = 10,000 or 20,000 have the smallest median OOB-ER as nTree > 10,000.

SFigures 5 and 6 show that OOB-ER is not affected by a combination of mTry and minNS since a lower mTry led to smaller OOB-ER for all minNS. While RFs with mTry of 0.2 and 0.5 with minNS of 10,000 resulted in comparable OOB-ER for all nTree, this is not true when minNS = 20,000 which shows clearer separation in OOB-ER between mTry when nTree > 15,000.

S3 **Hyperparameter Optimisation for VariantSpark with TOPMed Data**

As with S1, hyperparameter optimisation was conducted in R/v4.2.1^1^ with the R library **ranger**^2^ using the QC’d TOPMed (TM) CAD cohort (number of samples = 11,326 and number of genetic variants = 8,533,119). Note that this is the not the final dataset used to run the validation analysis which included the subset of genetic variants included in the UK Biobank CAD cohort (i.e., number of genetic variants = 3,996,295). However, the effect of this should be minimal since RFs are relatively robust against hyperparameters.

Again, batches of random forests (RFs) with various hyperparameter settings described below were built allowing for bootstrapping of the model metric OOB-ER to be performed and the stability of the metric across the hyperparameters can be evaluated. The following three key hyperparameters were optimised:

1. nTree: Number of trees in the forest
2. mTry: *Fraction* of total number of variants/features evaluated at each node of a tree
3. minNS: Minimum number of observations in a node to be processed.

The following sections (S2.1, S2.2, S2.3, and S2.4) will show that using OOB-ER, these following values are optimised for each hyperparameter:

- nTree = 10,000 – 15,000
- minNS = 1,000
- mTry = 0.1*nV
  S3.1 **nTree**

See S1.1 for a description of nTree.

A total of 250 batches of RFs were built with the following parameters:

- nTree = 100
- mTry = 0.1*nV (853,311)
- minNS = 10,000

Therefore, when the batches were joined, the cumulative number of trees evaluated is 25,000.

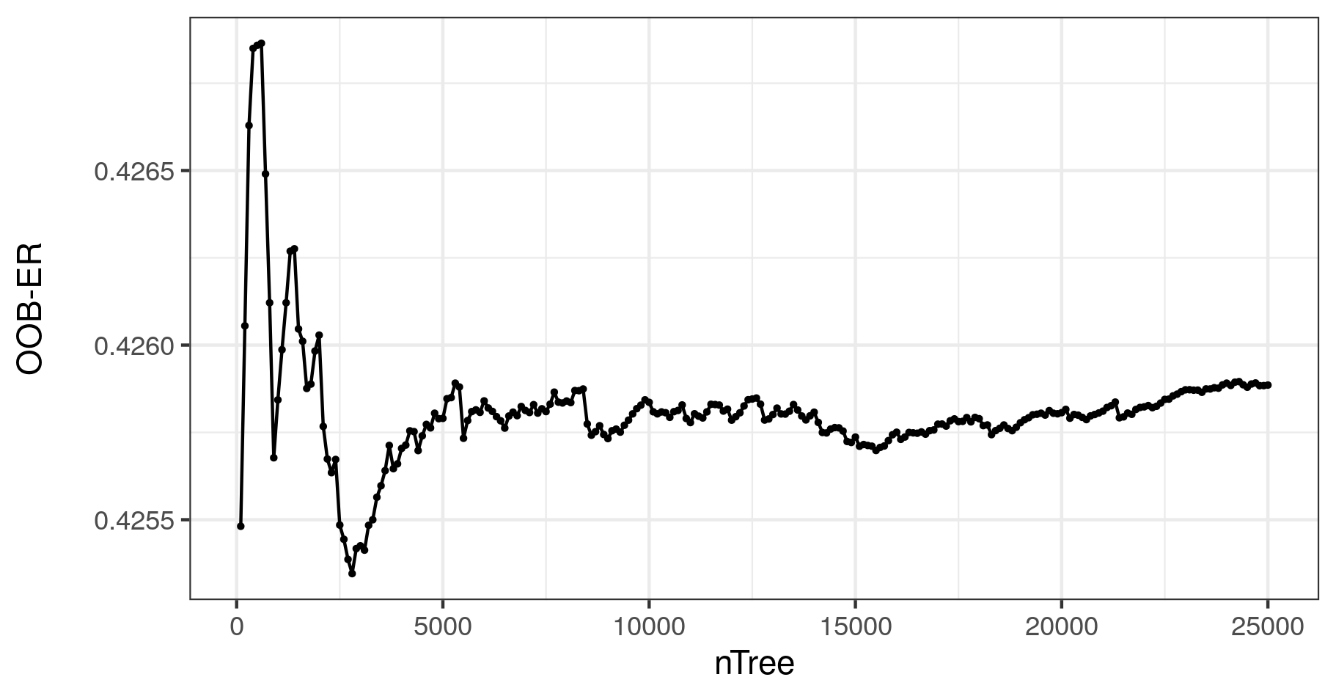

SFigure 8. nTree vs OOB-ER with TM data. As expected, OOB-ER is very variable when nTree is small (i.e., < 2,500) but starts to stabilise when nTree ≈ 5,000. The figure suggests that increasing nTree over 10,000-15,000 would result in more computational load for limited gains in OOB-ER.

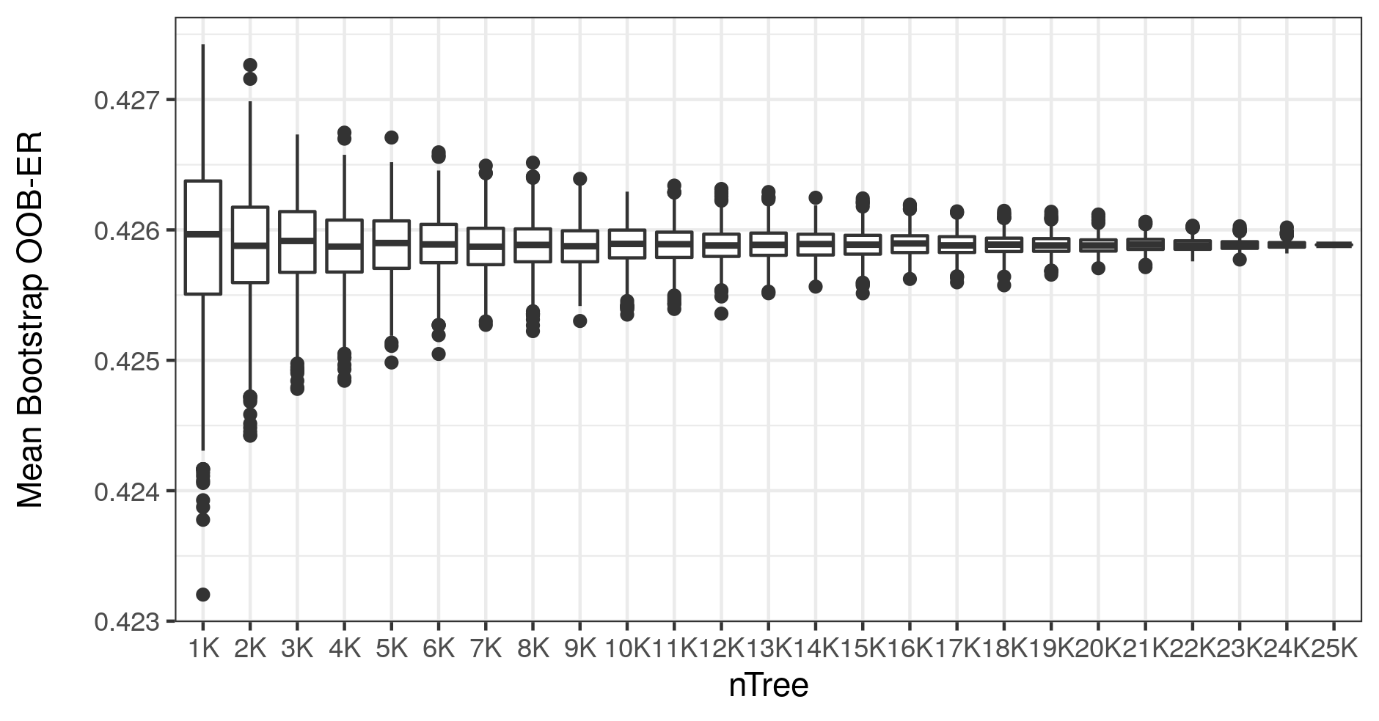

SFigure 9. nTree vs Bootstrapped OOB-ER with TM data. In concordance with SFigure 7, the variability of OOB-ER is large when nTree is smaller (e.g., < 3,000) but starts to become comparable as it gets larger. The distribution of OOB-ER across bootstraps are similar when nTree ~ 6,000 – 12,000) suggesting limited gains in building more trees between 6,000 – 12,000 trees.

As expected, OOB-ER decreases and stabilises as nTree increases (SFigures 7 and 8). Both figures suggest that the point of diminishing returns (i.e., when the decrease in OOB-ER no longer justifies increase in nTree) is when nTree = 10,000.

S3.2 **Minimum Node Size (minNS)**

See S1.2 for a description of minNS.

A total of 250 batches of RFs were built with the following parameters:

- nTree = 100
- mTry = 0.1*nV (853,311)
- minNS = 5,000, 10,000, 20,000

Therefore, when the batches were joined, the cumulative number of trees evaluated is 25,000.

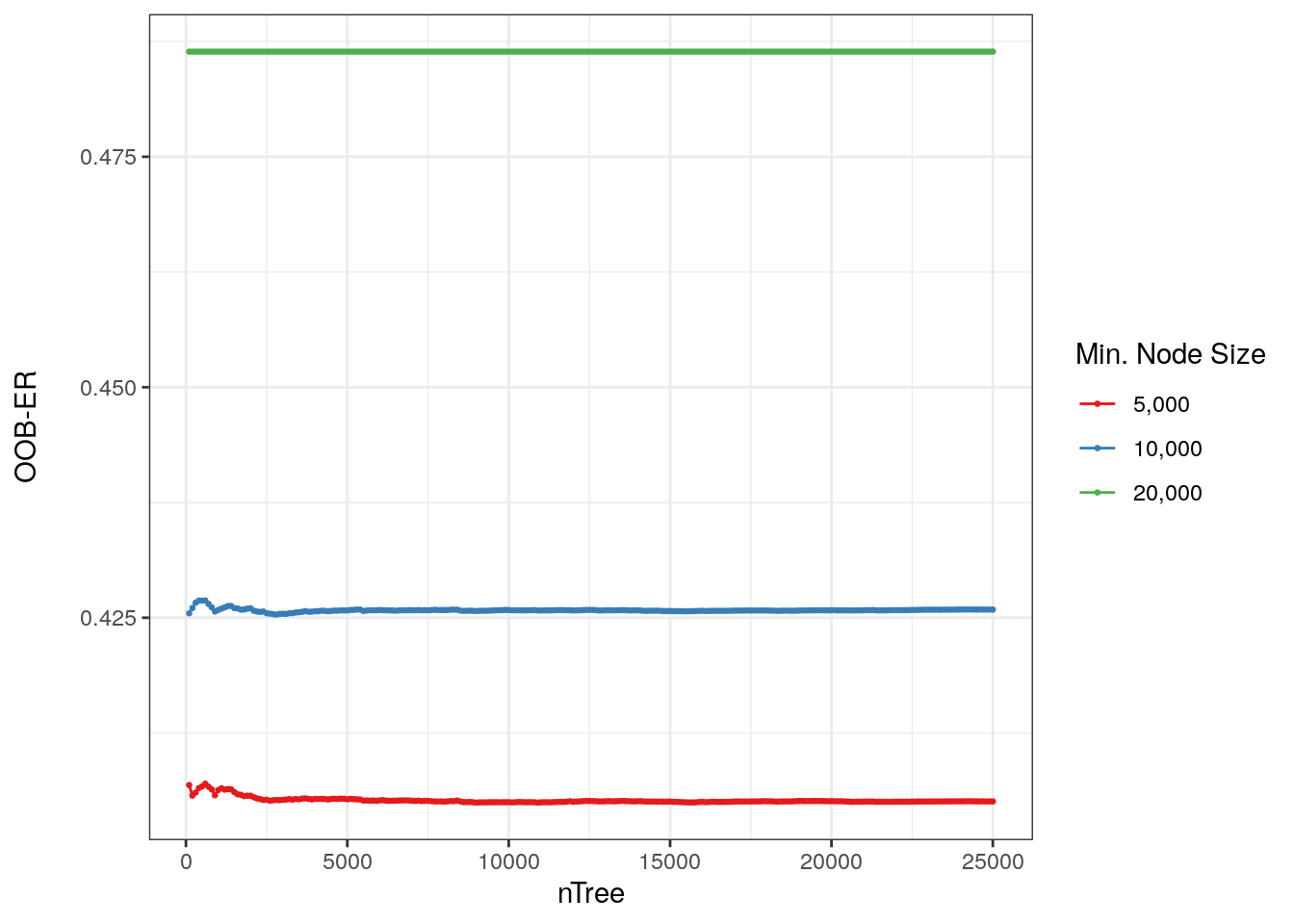

SFigure 10. OOB-ER vs nTree for various minNS with TM data. The figure shows a large decrease in OOB-ER when minNS decreases across all nTree. This is unlike the UK Biobank (SFigure 3). OOB-ER is clearly smallest when minNS = 5,000.

From SFigure 9, the minNS of 5,000 resulted in the lowest OOB-ER across all nTrees. Interestingly, this result is in opposition to that from the UKB dataset which suggested that RFs with higher minNS would be more accurate (S1.2). Furthermore, the range of OOB-ER from the TOPMed data is much wider than that of the UK Biobank data (range = 0.4 - 0.475 vs range = 0.466 - 0.471 respectively). There is some decrease in OOB-ER as nTree increases for minNS = 10,000 & 5,000 but not substantially, and as expected, OOB-ER stabilises when nTree > 5,000.

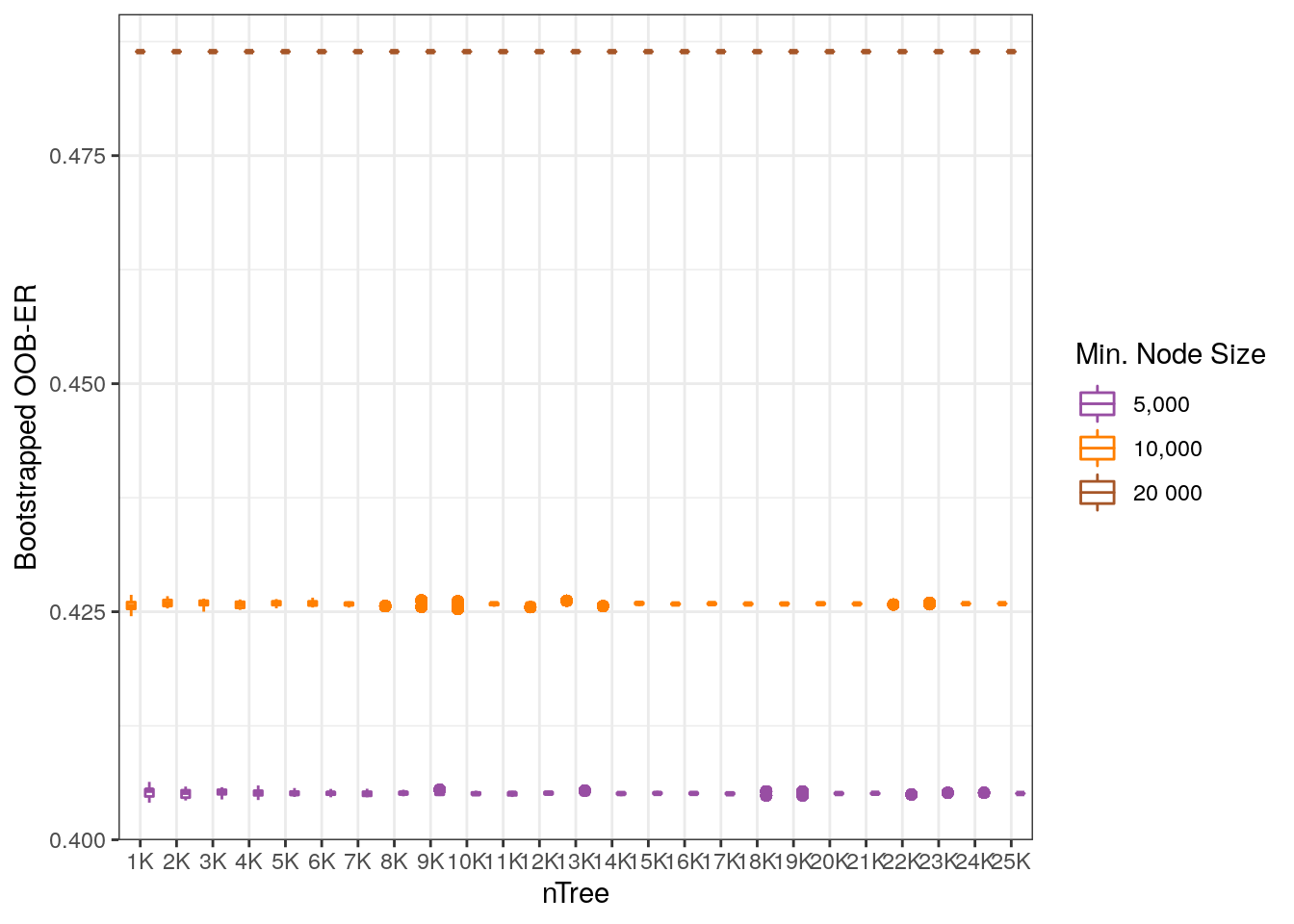

SFigure 11. Bootstrapped OOB-ER vs nTree for various minNS with TM data. As expected, given SFigure 9, there is clear separation of boxplots across minNS and nTree. The figure concurs that a minNS of 5,000 results in the lowest OOB-ER.

Unlike with the UKB data (SFigure3), there is no overlap of boxplots with the TM dataset with considering bootstrapped OOB-ER (SFigure 10). A minNS of 5,000 clearly results in the smallest OOB-ER across nTree for the TM dataset.

S3.3 **mTry**

See S1.3 for a description of mTry.

A total of 250 batches of RFs were built with the following parameters:

- nTree = 100
- mTry = (nV)^1/2^ (2921), 0.1*nV (853,311), 0.2*nV (1,706,622).
- minNS = 5,000, 10,000, 20,000

Therefore, when the batches were joined, the cumulative number of trees evaluated is 25,000.

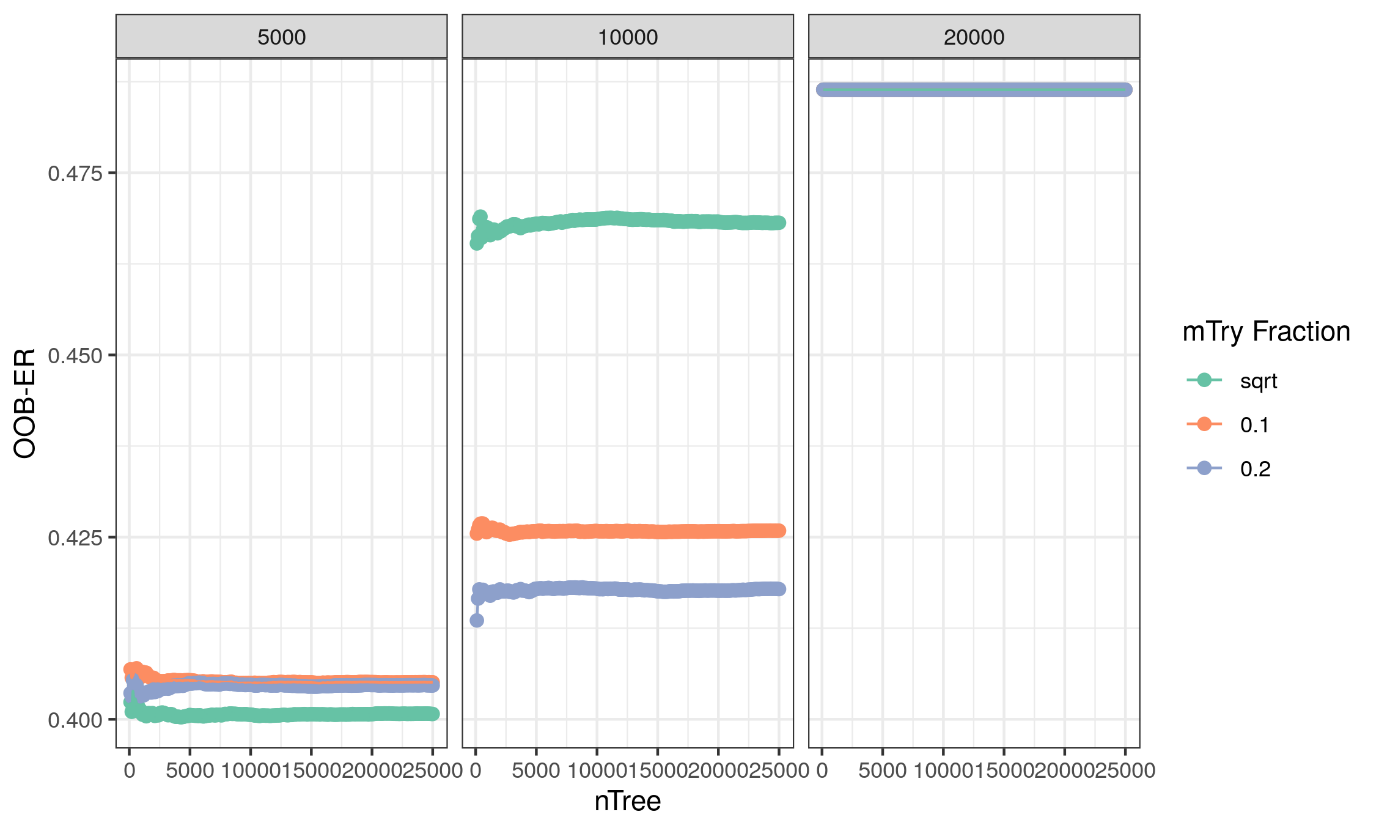

SFigure 12. nTree vs OOB-ER for various mTry facetted by minNS with TM data. There seems to be a combinatorial effect of mTry and nTree on OOB-ER as higher mTry when minNS is 10,000 leads to lower OOB-ER, but this pattern is not true when minNS is 5,000. For the test hyperparameters, the lowest OOB-ER resulted from the RF with mTry = (nV)^1/2^ and minNs = 5,000.

Due to the larger number of variants in the TM dataset, a mTry fraction of square root (sqrt) was used instead of 0.5 to reduce computational complexity when building the RFs. For all mTry fractions, a minNS of 5,000 still resulted in the lowest OOB-ER. When minNS = 5,000, a square root for the mTry fraction resulted in a marginally smaller OOB-ER.

S3.4 **More minNS**

Given that there was such a large difference in OOB-ER between minNS, which is also unlike the results from the UKB data, more minNS was trailed with two mTry fractions: sqrt and 0.1%.

A total of 250 batches of RFs were built with the following parameters:

- nTree = 100
- mTry = (nV)^1/2^ (2921), 0.1*nV (853,311)
- minNS = 1,000, 3,000, 5,000, 10,000, 20,000

Therefore, when the batches were joined, the cumulative number of trees evaluated is 25,000.

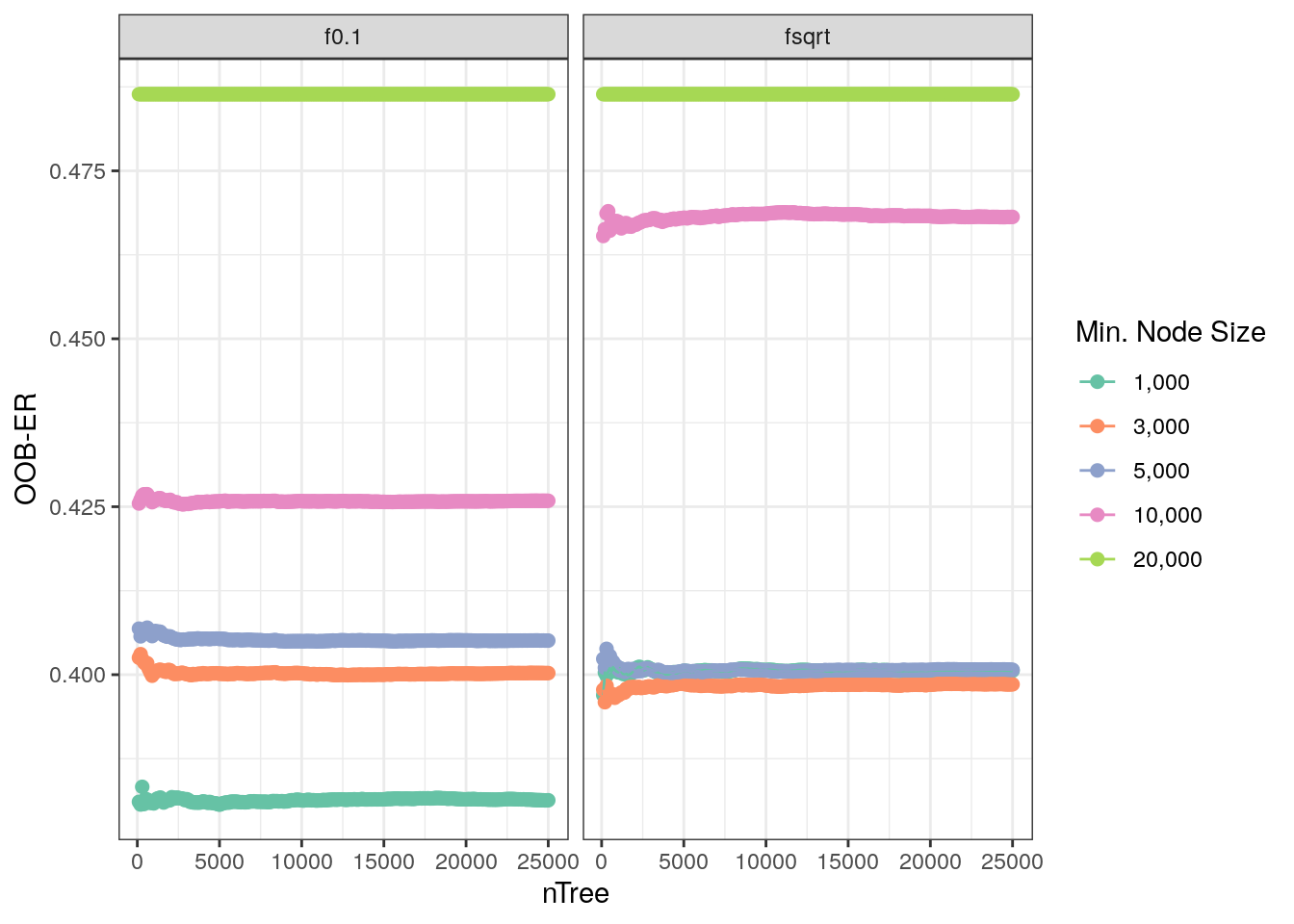

SFigure 13. nTree vs OOB-ER for various minNS facetted by mTry with TM data. There is a combinatorial effect occurring between minNS and mTry on OOB-ER such that OOB-ER is comparable when minNS = 3,000 & 5,000 for both mTry fractions, but not when minNS is larger at 10,000 and 20,000. For the tested parameters, the lowest OOB-ER resulted from the RF with mTry = 0.1*nV (853,311) and minNS = 1,000.

While results from SFigure 11 suggested a combination of mTry fraction = sqrt and minNS = 5,000 for the lowest OOB-ER, SFigure 12 shows a different set of hyperparameters for even smaller OOB-ER. When mTry fraction is set to sqrt, there is no discernible difference in OOB-ER when minNS = 1, 000, 3,000, and 5,000. However, when mTry fraction is 0.1 in combination with minNS = 1,000, the RF resulted in a considerably smaller OOB-ER.

For all combinations, OOB-ER stabilises when nTree > 5,000.
